# The State of Play of Parkinson’s Disease in Africa: A Systematic Review and Point of View

**DOI:** 10.1101/2023.07.07.23292392

**Authors:** Olaitan Okunoye, Yared Z. Zewde, Jihan Azar, Biniyam A. Ayele, Saiesha Dindayal, Ahmed Moustafa, Mosab Abdulkarim, Funmilola T. Taiwo, Daniel Gams Massi, Mohamed Salama, Abenet T. Mengesha, Yohannes D. Gelan, Dereje M. Oda, Fikru T. Kelemu, Charles Achoru, Vida Obese, Zeinab Kone, Momodou Cham, Maouly Fall, Abdoulaye Bocoum, Foksouna Sakadi, Philip Adebayo, Eric Guemekane Bila Lamou, Lorraine Chishimba, Aiysha Chaudhry, Ali Shalash, Eman Hamid, Musa Watila, Omotola Thomas, Oluwadamilola Ojo, Njideka U. Okubadejo, Mie Rizig, the International Parkinson’s Disease Genomics Consortium Africa (IPDGC-Africa).

**Affiliations:** UCL Queen Square Institute of Neurology, University College London, Queen Square, London WC1N 3BG, UK; Department of Neurology, School of Medicine, College of Health Sciences, Addis Ababa University, Addis Ababa, Ethiopia; Institute of Global Health and Human Ecology, The American University in Cairo (AUC), New Cairo 11835, Egypt; Division of Neurology, Department of Neurosciences, Faculty of Health Sciences, University of the Witwatersrand, Johannesburg, South Africa; Department of Biology, Systems Genomics Laboratory, American University in Cairo, New Cairo, Egypt; Nuffield Department of Clinical Neurosciences, University of Oxford, Oxford OX3 9DU, United Kingdom; Faculty of Medical Laboratory Sciences, Al-Neelain University, Khartoum, Sudan; University College Hospital, Ibadan; Douala General Hospital, University of Buea, Cameroon; Faculty of Medicine, Mansoura University, Mansoura, Egypt; Department of Internal Medicine, Jos University Teaching Hospital, Nigeria; Komfo Anokye Teaching Hospital, Ghana; Université des Sciences des Techniques et des Technologies de Bamako, USTTB, Mali; Richard Novati Catholic Hospital, Sogakope, Volta Region, Ghana; Service de Neurologie, CHN de Pikine, Dakar, Senegal; Department of Neurology, National Reference Teaching Hospital of N’Djamena, N’Djamena, Chad; Aga-Khan University, Medical College East Africa, Faculty of Post-graduate Medical Education, Dar es Salaam, Tanzania; Neurology Unit, Douala Gyneaco-Obstetric and Pediatric Hospital, Douala University, Cameroon; Department of Internal Medicine, University of Zambia, School of Medicine, Lusaka, Zambia; Department of Neurology, Faculty of Medicine, Ain Shams University, Cairo, Egypt; Department of Department of Neurology, Aberdeen Royal Infirmary, Forester hill, Aberdeen AB25 2ZN, UK; Parkinson’s Africa (Trustee Chair), 12 Constance St., International House London, E16 2DQ; Department of Medicine, College of Medicine, University of Lagos, Lagos State, Nigeria

**Keywords:** Parkinson’s disease, Africa, Education, Genetics, Management, Epidemiology, Patient, Risk Factors, Clinical Features

## Abstract

**Introduction:** Parkinson’s disease (PD) has become a global public health challenge as disability and death due to the disease are growing rapidly in comparison to other neurological disorders. There are no up-to-date comprehensive reviews on the epidemiology, environmental and genetic risk factors, phenotypic characterization, and patient-reported outcomes of PD in Africa. This data is crucial to understanding the current and future burden and suggesting actionable and/or researchable gaps aimed at improving disease outcomes.

**Methodology:** We conducted a systematic literature search using the electronic databases of Cochrane Central Register of Controlled Trials (CCRT), EMBASE, Medline, PsychINFO, Web of Science, Cumulative Index to Nursing and Allied Health Literature (CINAHL), African Journals (AJOL) and other unpublished literature. We included all studies providing data on people with PD in Africa from the start of each database till February 2023. Studies were not restricted based on diagnostic criteria or language. Outcomes of interest were summarised based on epidemiology, genetics, environmental risk factors, clinical characteristics, patient-reported outcomes (experience and quality of life), disease management and outcomes, access to care, patient support, and healthcare workforce training. We also investigated collaboration between African countries (internal) and across continents/world regions (external) and journal impact factors.

**Results:** A total of 4,855 articles were identified, of which 180 were included in this review. The majority were published from North Africa (mainly from Tunisia, and involved collaboration with investigators from France, the United Kingdom, and the United States of America). West Africa (Nigeria), Southern Africa (South Africa) and East Africa (mainly Tanzania) also had a relatively high number of publications. Methodological design varied across studies. Based on the pre-determined outcomes, articles identified were genetics (67), clinical features (65), environmental risk factors (16), epidemiology (14), patient experience and quality of life (10), management and access to care (5) and education and training (3).

**Conclusions:** The main hubs of PD-related research output in Africa are the Northern, Western and Southern regions of Africa (although with limited involvement of countries within these regions). External collaboration (outside the continent) currently predominates. There are considerable actionable and researchable gaps across all outcomes of interest, with a dearth of published information on health workforce capacity building, disease management and access to care, patient and caregiver engagement, and quality of life of people with PD in Africa. We recommend strengthening existing and emerging intercontinental networks for research, education, training and policy formulation and funding, leveraging on more recent developments such as the International Parkinson’s Disease Genomics Consortium-Africa (IPDGC-Africa), the International Parkinson and Movement Disorder Society Africa Section (MDS-AS), World Health Organisation (WHO) and initiatives with similar objectives.

## Introduction

Parkinson’s disease (PD) is the fastest-growing neurological disorder in terms of prevalence, disability, and deaths according to the recent Global Burden of Disease (GBD) data^1, 2^. A recent report by the World Health Organisation (WHO) declared PD a “public health priority”, stating that an immediate public health response is required to address the lack of resources required to manage the disease particularly in low- and middle-income countries (LMICs)^3, 4^.

Globally, PD is estimated to affect 1.5 to 26 per 100,000 of the general ageing population and the burden is projected to continue to increase by more than double in the next two to three decades, with the greatest impact expected in Africa^2, 5–8^. Africa is the second largest and second most populous continent, comprising 5 regions and 56 countries and dependencies (Eastern Africa (18), Western Africa (17), Central Africa (9), Northern Africa (7) and Southern Africa (5)) and a population of approximately 1.3 billion^9^. The proportion of the African population aged >60 in 2020 was 4.9%, with a resultant population at risk of PD (based on the peak age at onset of PD) of more than 63 million^10^. Despite this high estimate, there are considerable gaps in the depth and breadth of our understanding of the profile and impact of PD in Africa.

PD affects all ethnic groups and despite the predicted increase in age-adjusted prevalence of PD, studies on the epidemiology of PD in Africa repeatedly report low prevalence and incidence rates compared to Europe and North America^2, 11–17^. Methodological differences (mainly case ascertainment), underpowered studies, use of crude rather than age-standardized data for comparison, and a general sparsity of high-quality studies are presumably inherent biases that underlie these data. Also, geographical, and environmental differences in exposure to risk factors, and genetic variability are plausible explanations^17–19^. The sparse data on various aspects of PD in Africa may also be attributed to prevalent under-diagnosis and under-reporting emanating from the relatively few numbers of the neurological workforce (0.1 per 100,000) with expertise in research, diagnosis, and management of PD and other neurological disorders^9, 20, 21^. The 2016 GBD systematic analysis of data focused on PD, highlights specific deficiencies in the availability of comparative data from Africa including a lack of mortality and survival data^8^. A comprehensive review of the current landscape of PD in Africa is crucial to document a more recent baseline for measuring advances and time trends, and provide evidence to highlight healthcare disparities and issues related to healthcare systems and service provision for people with PD.

This systematic review aims to provide a summary of available information on epidemiology, genetics, environmental risk factors, clinical characteristics, patient-reported outcomes (experience and quality of life), disease management, access to care, patient support and healthcare workforce training of PD in Africa. We have not collected data on laboratory studies as this is outside the scope of this review. We also provide some recommendations to help with improving PD research in Africa. In addition, this review will guide researchers and policymakers in developing and prioritizing projects and programs aimed at addressing unmet needs in PD research and improving the health outcomes of people with PD^22^.

## Methodology

This study followed the Preferred Reporting Items for Systematic Reviews and Meta-analysis (PRISMA) guidelines for the search, extraction, synthesis of results and reporting^23^. Details of search strategy, inclusion and exclusion criteria, study selection criteria and data extraction and methodological quality assessment are included in supplementary material 1.

The searches were conducted in four stages between May and June 2020 and updated in February 2023. The first search was conducted systematically in 7 electronic databases including MEDLINE and EMBASE using OVID interphase, Web of Science, Science Citation Index Expanded (SCIE), PsycINFO, Cochrane Central Register of Controlled Trials (CCRCT), Database of Abstracts of Reviews of Effects (DARE) and Cumulative Index to Nursing and Allied Health Literature (CINAHL) using Medical Subject Headings (MESH) and keyword terms where appropriate. This search was initially conducted in Medline (Appendix 1) and then in other databases. The second search was conducted electronically in African Journals and in some local databases. Some of the local searches were conducted manually. The third search was conducted by searching internet resources and theses. The fourth search involved informal consultations through electronic mails and telephonic discussions with local neurologists for information on PD in Africa. In addition, we searched for potentially useful references cited in the key articles and available review articles. Original studies with any design were considered potentially eligible, if the participants included were African, had PD, and if the studies provided information on any of the focus areas including epidemiology, genetics, environmental risk factors, clinical characteristics, patient-reported experience and quality of life, disease management and outcomes, access to care, patient support, and healthcare workforce training. There were no restrictions regarding PD diagnostic criteria, language, or year of publication. Expert opinions, some letters to the editor, case reports, editorials, and reviews were excluded. A standardized data extraction form was used to record data of interests and the Newcastle-Ottawa scale was used to conduct the quality appraisal of included studies^24^ (Supplementary material 1 and Appendix 2).

## Results

The search of electronic databases as shown in the PRISMA diagram (Figure 1) identified 4,855 studies. After removing duplicates, 3,442 records identified by the systematic search were screened. Twenty-nine more articles were identified from local search (manually within African countries) and other sources. Potential full-text articles from 429 studies were then assessed for eligibility, and of these, 278 articles were excluded with reasons as shown in Figure 1. Finally, we captured data from a total of 180 articles (Figure 1).

**Figure 1:**
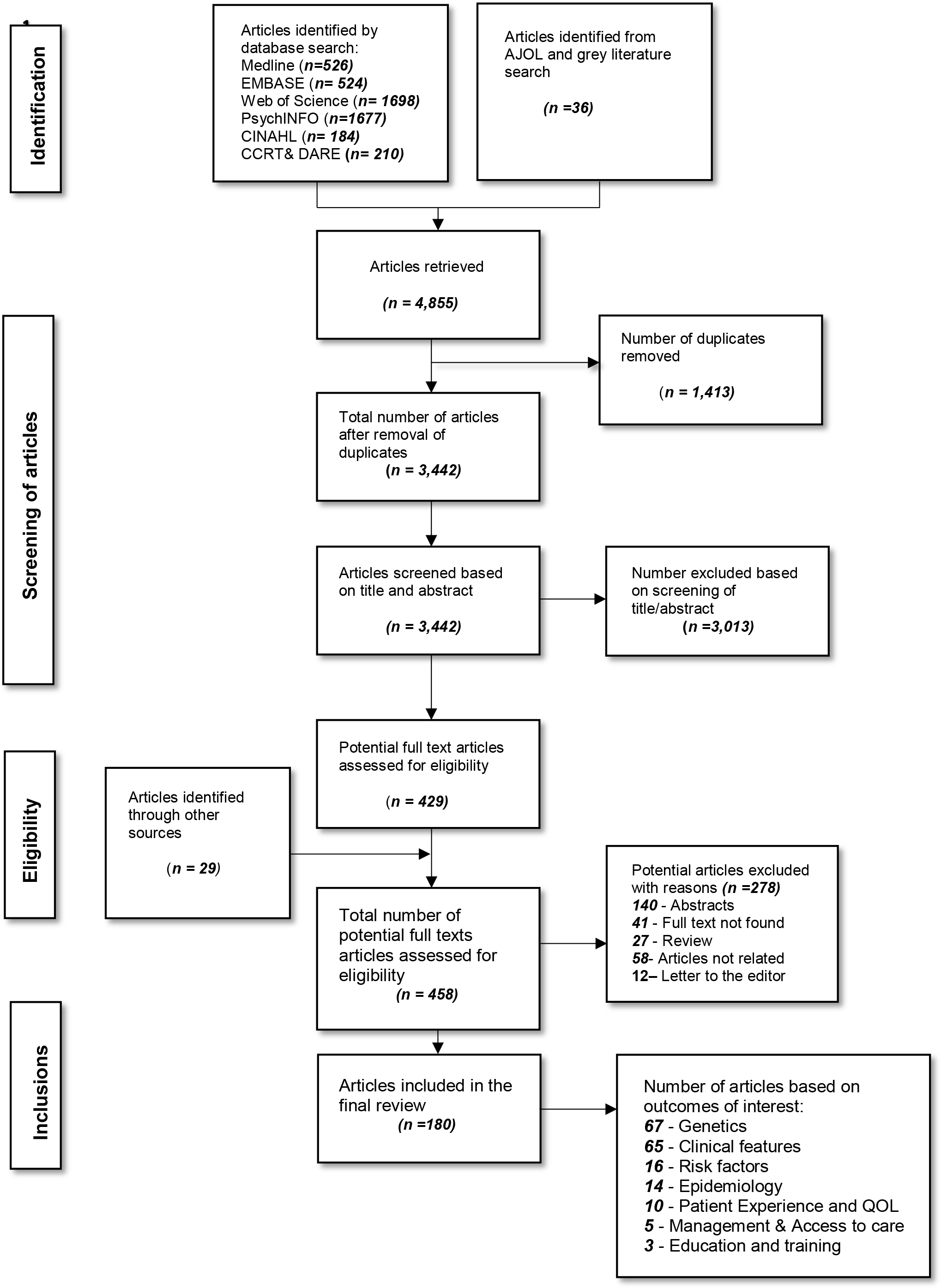
Diagram of the selection procedure to identify articles included in the systematic review.

The 180 studies identified in this review were conducted in 20 countries out of the 54 African countries. 68 (38%) were conducted in North Africa, 35 (19%) in West Africa, 34 (19%) in Southern Africa, 28 (16%) in East Africa, 7(4%) from Central Africa (all from Cameroon) and 8 (4%) other studies were from collaborative efforts between African countries (Table 1, Figure 2 and Supplementary Tables 1 and 2).

**Figure 2:**
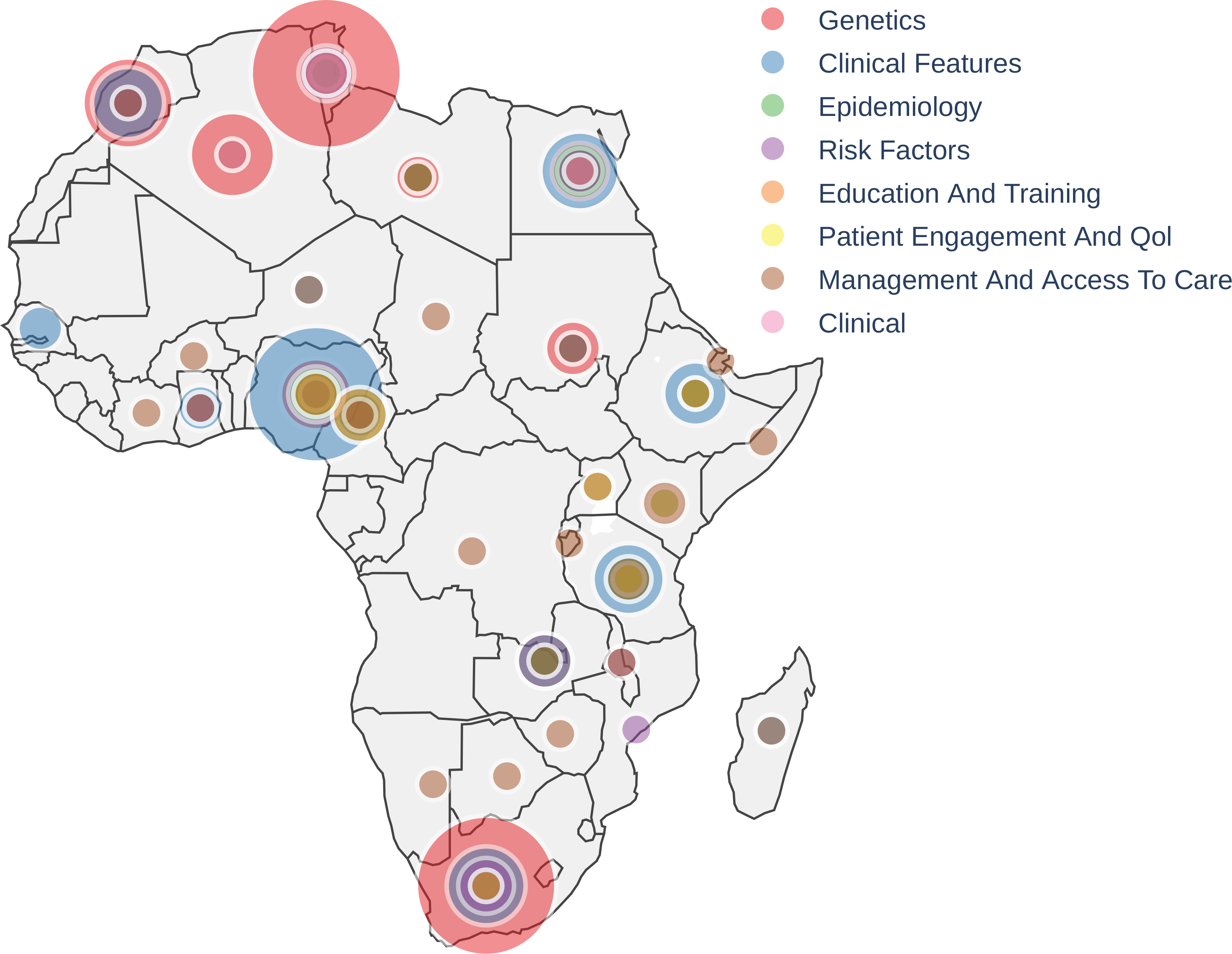
Outcomes of interest by African Regions/Countries.

**Table 1:**
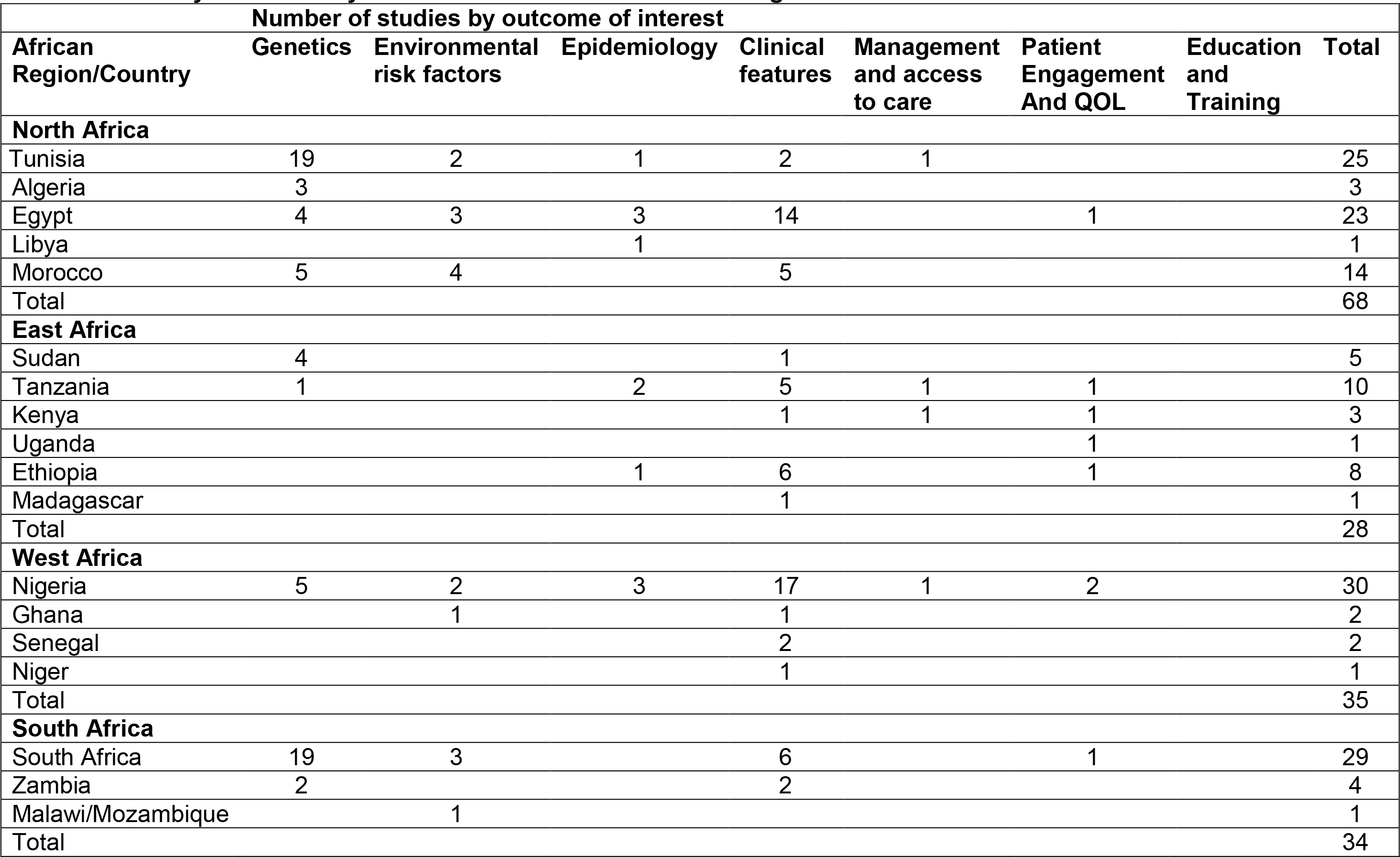
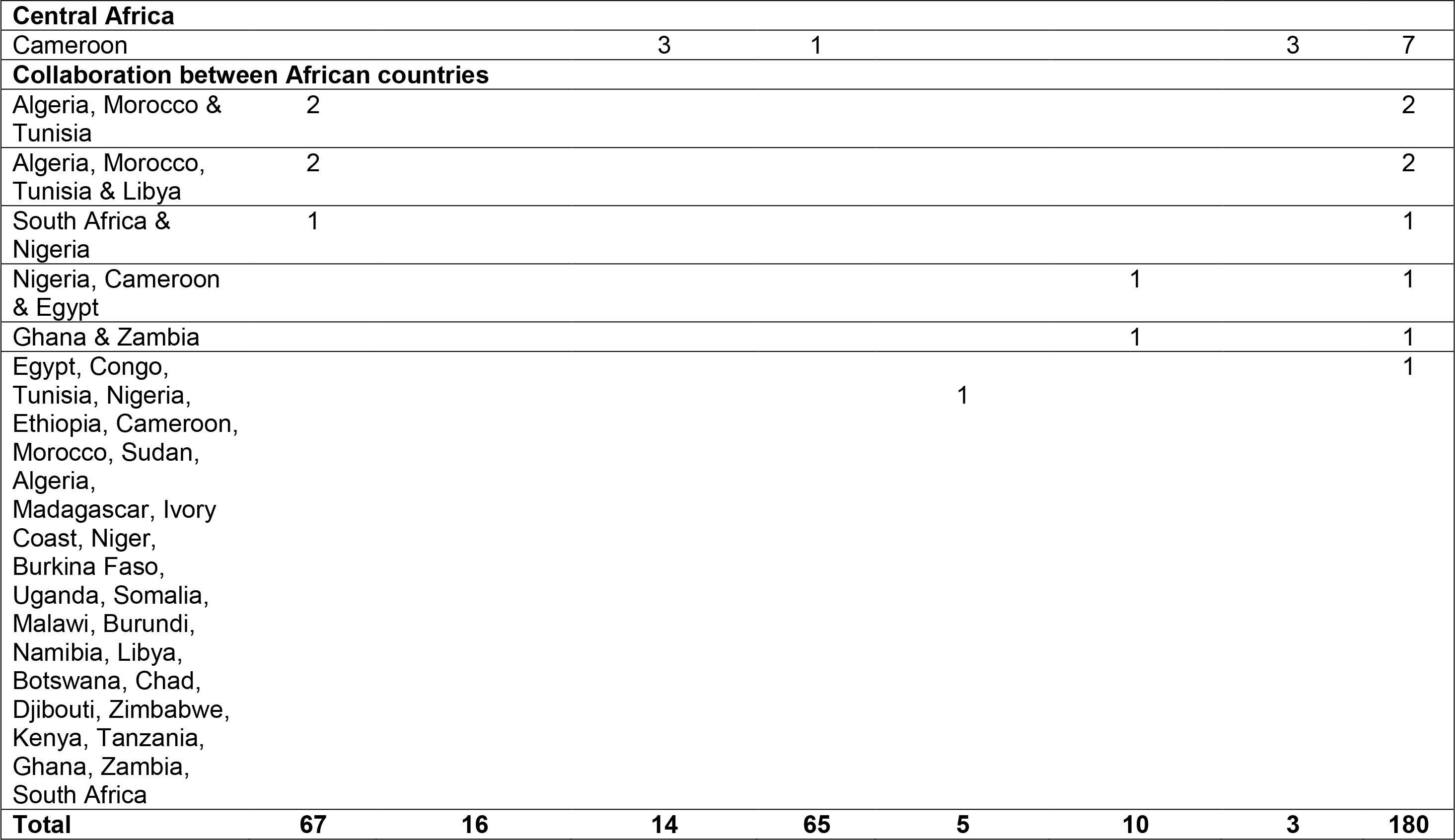
Summary of studies by outcomes of interest and African regions and countries.

The driving hubs of PD research in Africa are Tunisia (with 25 articles) and Egypt (with 23 articles) in North Africa, Nigeria (with 30 articles) in West Africa and South Africa (with 29 articles) in Southern Africa. There were no studies identified from the remaining 34 countries-Angola, Côte D’ivoire, Burkina Faso, Mali, Chad, Somalia, Djibouti, South Sudan, Zimbabwe, Guinea, Rwanda, Benin, Burundi, Togo, Sierra Leone, Congo, Liberia, Central African Republic, Mauritania, Eritrea, Namibia, Gambia, Botswana, Gabon, Lesotho, Guinea Bissau, Equatorial Guinea, Mauritius, Eswatini, Comoros, Cabo Verde, DRC, Sao Tome & Principe and Seychelles.

Observing trends of the number of studies per year showed that publication of studies slowly increased from 1998 to 2022, with around 90% published after 2010. The highest rise in the number of studies was noticed in the Northern region of Africa with studies mainly in genetics. This was followed by West Africa with most studies in clinical features and Central Africa where studies on education and training remained static (Table 1, Figure 3 and Supplementary Table 1).

**Figure 3:**
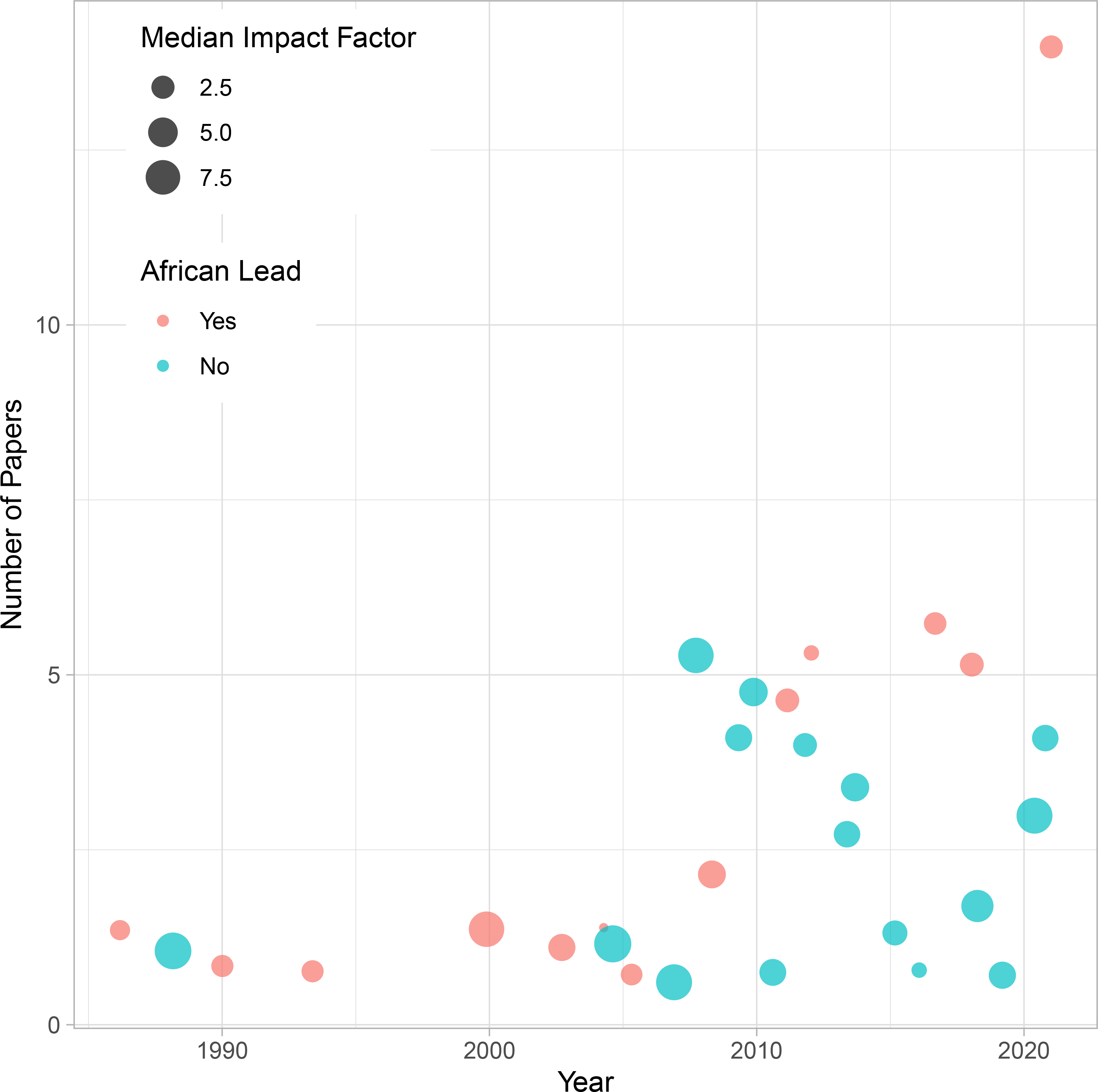
Number of Articles and IF per year.

The outcome of interest with the highest number of studies was the genetics of PD in Africa with 67 articles^25–91^. Clinical characteristics/features were described in 65^92–156^, 14 reported epidemiology^14, 157–169^, 16 studies investigated environmental risk factors of PD^81, 170–182^, and 10 studies examined patient experiences and quality of life (QoL)^183–192^. Studies which examined management and access to care (6)^20, 193–197^ and education and training (3)^198–200^ were the least (Table 1, Figure 2 and Supplementary Tables 1 and 2).

Approximately 73% of the included studies were cross-sectional in design with many including unaffected controls (Supplementary Table 1). Longitudinal and registry-based studies were not identified. All the studies identified through our search were in English except for 1 study from Senegal which when translated by one of the French speaking authors was an abstract and was not included in the review. Around 60% of the included studies had medium quality based on the Newcastle Ottawa scale scoring. We did not exclude studies based on quality due to the paucity of data in Africa.

We collected data on research funding and collaboration within African countries and internationally. This data was derived from reviewing the author lists, affiliations, and sources of funding as reported by the authors. This showed that collaboration and funding were available in 82 (45%) of the retrieved studies, mostly external, with the United Kingdom, the United States of America and France being the main collaborators. Collaborations between African countries were reported between Algeria, Morocco and Tunisia; Nigeria, Cameroon and Egypt; South Africa and Nigeria; and Ghana and Zambia (Table 1 and Supplementary Table 1).

African region which reported most funding was Northern Africa mainly Tunisia with funding from the United Kingdom, USA and Europe. This was largely driven by genetic studies investigating LRRK2 in the region. Western Africa and Central Africa however received less funding. Funding from African organisations remains scarce and reported in 35 studies. Of note is that South African studies in comparison to other countries were mostly funded by local South African organisations.

We also observed that the Impact factor (IF) of Journals where African authors published their research ranged from 0.03 (International journal of pharmaceutical research in Egypt) to 44.1 (lancet neurology in Tunisia) with most articles which were led by non-Africans published in Journals with the highest IF. Of the 180 studies included, 154 studies had African researchers as leading authors.

### Genetics of Parkinson’s disease

Sixty-seven articles investigated the genetics of PD in Africans living in 10 African countries (Tunisia, Algeria, Egypt, Morocco, Sudan, Tanzania, Nigeria, South Africa, Zambia and Libya) with around 77% of these conducted in Northern (mainly Tunisia) and Southern Africa (mainly South Africa). There were no genetics studies conducted in populations living in Central, Eastern and the French speaking West coast (Table 1, Figure 1 and Supplementary Table 1). Forty-six percent of these genetic studies applied targeted genotyping to screen for causal variants in PD known genes^26, 28, 30, 31, 33–45, 47, 48, 51–58, 62–66, 68, 69, 71–78, 80, 81, 84, 86^. Around 19% most of which were from Tunisia, investigated phenotypic-genotypic correlations^25–27, 29, 32, 33, 39, 50, 60, 68, 70, 78, 82^. Others looked at pattern of inheritance^25, 27, 30, 35, 59, 79^ ancestral dating of mutations^46, 67^ and functional analysis^61, 78, 85^.

The most represented ethnicity was the Arab-Berbers in North Africa.^31, 32, 35, 40, 43, 59, 67, 79^. Study populations varied between familial populations (30.6%) ^25–31^ ^35–37, 39, 47, 53, 54, 60, 72, 77, 79, 85^, sporadic (22.6%)^32, 46, 49–51, 57, 61–63, 78, 79, 81–83^, mixed (35.5%)^33, 34, 38, 40–46, 48, 56, 58, 59, 65, 68–70, 73, 75, 76, 80^ and not otherwise reported (11.3%)^33, 55, 64, 66, 67, 71, 84^. Positive mutational analysis was more likely in familial cohorts^26, 28, 30, 35, 37, 47, 53, 63, 72, 77, 86^. Cohort sizes were modest ranging between 8 in Sudan and 778 in Tunisia (Supplementary Table 1).

The commonly screened genes were *LRRK2, PARK2, PINK1, SNCA, DJ-1, ATP13A, MAPT, EIF4G* and *VPS35*, which were reported as the mendelian genes associated with PD. Of these genes, *LRRK2* was the most frequently investigated, particularly in North Africa (Tunisia, Algeria, Morocco, and Egypt)^28, 29, 31–35, 39, 41–43, 45, 46, 49–51, 54, 55, 59, 60, 62, 64, 67–70, 73, 75, 79, 80, 83, 84, 86^.

In addition, the *LRRK2 G2019S* mutation was the most frequently reported, with a frequency ranging from 1.45% to 43%^28, 29, 31–33, 35, 41, 43, 49–51, 53, 59, 60, 62, 64, 67–70, 73, 75, 79^. These studies included cohorts which were familial, ^28, 29, 31, 35, 39, 54, 60^ sporadic,^32, 49–51, 79, 83^ mixed,^33, 34, 41–43, 45, 46, 59, 62, 66, 68, 69, 73, 75, 80^ or not stated^51, 55, 64, 67, 84^. It isnoteworthy that some of these studies included individuals who were previously accounted for in prior studies^25, 26, 31, 32, 93^. The highest frequency of *LRRK2 G2019S* globally was reported in Morocco (41%) in a cohort that included familial and sporadic PD^68^.

*LRRK2 G2019S* has also been investigated in South Africa, Nigeria and Zambia with no mutations reported in black sub-Saharan Africans^41, 54, 84^. Around 16 studies investigated the clinical features in *LRRK2* carriers from North Africa^25–27, 31, 39, 42, 49–51, 59, 60, 64, 68, 70, 74, 201^. At the time of this review, there were no studies which used whole genome sequencing or genome-wide association approaches. In addition, we did not identify studies which included multiple ancestral groups from multiple African regions in genetic studies.

### Environmental risk factors for PD

Our review identified 16 studies which reported data on environmental risk factors for PD in Africa. Of these, 9 were from Northern Africa (Tunisia, Morocco and Egypt), 4 from Southern Africa (South Africa and Mozambique) and 3 from Western Africa (Nigeria and Ghana). The authors did not report any prior reviews to guide the pre-selection of the putative risk factors.

These studies focused on three major categories of environmental risk factors including exposure to heavy metals, infections/comorbidities and gene-environment interaction. Three studies (from Tunisia and Egypt) investigated gene-environment interaction among Africans with PD,^81, 174, 177^ four studies (from South Africa, Morocco and Nigeria) investigated exposure to pesticides, heavy metals (manganese)^175, 176, 178^, and similar trace elements^172^. These studies showed a positive association of these risk factors in the development of PD, but most were small studies with plans for future large-scale studies.

The link between typhoid and hepatic disorders and the development of PD was investigated by two studies from Malawi-Mozambique and Egypt^170, 173^. One study (from South Africa) examined the risk of PD and demographic characteristics such as ethnicity and family history^171^. Another study from Ghana investigated nutritional status and dietary habits in a pilot study^197^. More recent studies have looked at multiple factors^182^ (such as consumption of well water, occupation, etc), immunological factors (for example tumour necrosis factor-a), serum uric acid and stool samples for gut micro-organisms^180–182, 202^. Most of these studies were under-powered and thus had inconclusive results requiring further large-scale investigations.

### Epidemiology

Our review showed that there is a dearth of data on epidemiology of PD in Africa. There are no available registries or national health record systems. Studies on the prevalence of PD in Africa have been population-based. The crude prevalence of PD was reported as ranging from 7 (Ethiopia having the lowest) to 436 (Tunisia having the highest) per 100,000 person-years^14, 157–160, 165, 167^ and increasing with age, from 452.1 per 100,000 in people aged 40 years and over^165^ to 590 per 100,000 in people aged ≥70 years^163^. Age-adjusted prevalence was reported in 4 studies with rates ranging from 40 to 68 per 100,000 person-years^14, 157, 158, 160^. Whilst three^14, 157, 158^ of these studies used indirect age standardization^14, 157, 158^, one used direct standardization^160^. Two studies reported a crude incidence of 4.5 per 100,000/year (Libya) in 1985 and 62 per 100,000/year in 2013 (Egypt)^157, 167^. Retrospective hospital-based studies conducted in Cameroon and Nigeria reported frequencies ranging from 0.79% to 10.2%^161, 162, 166, 168^.

### Clinical features

The clinical features of PD in Africans were examined in 65 studies with 35% from Western Africa (mainly Nigeria with 17 studies), 32% from Northern Africa (mainly Egypt with 14 studies), 21% from East Africa (mainly Tanzania with 5 studies), 12% from Southern Africa (mainly South Africa with 6 studies) and 1 study from Central Africa (Cameroon)^92–156^. The mean age of people with PD was reported to be between 50 and 60 years, and there was a male preponderance with most studies reporting male: female ratio of around 2.5-3:1^92, 99, 112^. However, one study reported a larger proportion of females in their participants with PD^119^.

The tremor-dominant phenotype was reported in 60% of the 65 studies^94, 111, 116^, with the classical rest tremor of the hand reported in 42.5% ^99, 129, 203^. Non-motor symptoms have also been the focus of some studies with neuropsychiatric features, gastrointestinal symptoms, and cognitive impairments being the most frequently reported^109, 125, 129^.

### Management and Access to care

Of the five studies (from Tunisia, Tanzania, Kenya, Nigeria and collaborative study of African Countries-Table 1) which investigated management and access to care, two studies conducted in Kenya and Nigeria examined the availability and affordability of medicines using the WHO/Health Access International methodology^194, 195^. Anticholinergics were reported to be the most available and accessible PD medication, dopamine receptor agonists were the second most available while levodopa formulations were the third most available medication^194, 195^. Anticholinergics and levodopa were reported to be more available in private pharmacy outlets than in public ones (80.8/4.8% versus 47.2/18.2% and 74.8/1.5% versus 13.2/ 9.2% respectively)^194, 195^. The long-term treatment, response to treatment and feasibility/sustainability of supplying PD medications to clients in rural settings were investigated by a study conducted in Tanzania^193^. The authors reported the follow-up of persons with PD on a levodopa/carbidopa combination for three years and found that it was possible to treat and follow up PD patients, but the availability of PD medications was a major challenge^193^. In addition, a recent continent-wide survey of the availability and affordability of PD therapies showed similar findings with low availability and affordability of most medications in most African countries^20^.

### Patient Engagement and QoL

Of the 10 articles that investigated patients’ experiences and quality of life (QoL) included in this review, 4 focused on the QoL of persons/people with PD in Nigeria. Three of these reported poor QoL, using the PDQ-39 in people with PD compared to age and sex-matched controls, and disease duration, severity and co-existing depression were reported as the main determinants of QoL^184, 185, 190^. Cognitive behavioural therapy was reported to improve QoL in PD in one study^189^.

Another study from Egypt reported poor QoL related to sexual dysfunction in PD, and this was worse in older male patients^192^.

The other five studies focused on patient and/or caregiver experiences in Tanzania, Uganda, Ethiopia, South Africa, and Kenya. These studies reported that people with PD in Africa have a limited level of knowledge of the disease and this is characterized by misconceptions and stigma surrounding the disease^183, 186–188, 191^. In addition, a lack of adequate support and resources for people with PD and their caregivers and feelings of poor quality of life were also reported^183, 186–188, 191^.

### Education and Training

The focus of one of the three studies on PD education and training from Central Africa (Cameroon) was to assess how much knowledge neurologists practising in Africa have on neurological diseases including movement disordors^198^. The authors reported that neurologists on the continent were more familiar with central nervous system (CNS) infections compared to movement disorders^198^. The other two studies focused on tele-education for PD using lectures, video conferences, and other lecture materials to improve the knowledge of PD and other movement disorders among healthcare workers and medical students^199, 200^. Both studies reported that in addition to having an elevated level of satisfaction following the program, participants recorded a remarkable improvement in their knowledge of PD through the pre- and post-course assessment^199, 200^.

## Discussion

Africa is the second largest populous continent constituting 17% of the world population. By the year 2050, this figure is expected to reach 26%. The average life expectancy across the continent is expected to rise by 20 years from 49 years to 62 years by the end of the century^2, 5–8^. This rapid rise in population size and age is expected to be accompanied by a steep increase in the burden of neurodegenerative diseases particularly PD. However, preparedness in the continent to deal with this foreseeable burden is lacking.

In this systematic review, we present a comprehensive summary on a wide scope of data on the PD landscape in Africa, including epidemiology, genetics and environmental risk factors, clinical profile, management, patient-related issues, health workforce capacity building and research collaboration. This systematic review updates earlier reviews and widens the scope of enquiry, providing additional insights into the current scenario and a basis for benchmarking future policies.

First, we report that PD African research output, despite a rise in numbers in the last ten years, remains poor, constituting around 0.001% of the total PD research publications globally. At the time of this review, published data emanated from studies conducted in less than 50% of the African countries located primarily in three of the five regions (Northern mainly Tunisia and Egypt, Western mainly Nigeria and Southern Africa mainly South Africa). Given the intra-continental diversity in geographical, socioeconomic, ethnocultural and genetic factors, expanding research output to ensure the representativeness of the entire continent is critical. This will address the impediment of non-generalizable findings and diminish barriers preventing un-researched populations from experiencing the benefits of innovations. In addition, the relevance of advances in preventative and therapeutic strategies would be better understood, and policies guiding the allocation of scarce resources would be sufficiently backed by data to prioritize and bring PD into cognizance across all African countries.

Second, our review found the focus of PD research in Africa has been on genetics, clinical profile/phenotypes, and epidemiology (prevalence studies and hospital frequency), with no large-scale clinical trials reported to date. We observed a trend towards region-specific ‘preferences’ in PD research focus, with a predilection towards studies on genetics prevailing in publications from Northern Africa, descriptions of clinical profiles from Western Africa (Nigeria), and a mix of clinical and genetics studies predominantly, from Southern Africa (South Africa). Data on environmental and other risk factors/biomarkers for PD, management, and access to care of people with PD, patient engagement and QoL, education and training are lacking from the continent. The research focus is probably driven by a combination of factors including research experience and research leadership, availability of infrastructure and resources (including funding) and technological support, competing interests, perceptions that determine the focus of industry-sponsored research, awareness of research gaps and prioritization of research agenda, and international collaboration.

Our review showed that research productivity paralleled the presence of international collaborations. Most studies with external collaboration got published in high-impact journals in comparison to those without. Also, most of the studies with African lead-authors were published in low-impact journals compared to those with an international lead-author. The impact factor appeared to increase with external (international) collaboration and when the lead author was non-African. Most collaborations that involved more than one country were with researchers in Europe (France and the United Kingdom) and North America (the United States of America). More expertise- and resource-intensive research was seen from regions with international collaboration. For instance, Northern and Southern Africa, the regions with greater access to research funding and external collaboration predominated in PD genetics output, whereas more low-cost clinical research was mostly conducted in West Africa^41, 49, 59, 70, 204, 205^.

Of note, much of the genetics research interest in North Africa, particularly Tunisia, has been prompted by the interest generated by the predominance of the *LRRK2 G2019S* founder mutation in the Northern African Arab and Berber population^32, 59, 204^. Majority of these studies have however been gene-targeted or replications with very few or no published studies focused on wider strategies for new gene discovery, and no whole genome or whole exome sequencing data to date.

Third, our review showed that the most employed study design was cross-sectional, with varying sample sizes leading to poor power and inconclusive results. This could partly be due to a dearth of epidemiological data which is the backbone of research and healthcare planning. Most African countries lack accurate and up-to-date census data, health records and systems where data can be collected for longitudinal studies and larger studies with more power. These are largely attributable to the lack of political wealth of most African countries.

Fourth, we report the paucity of neurology education and training, which continues to be a challenging problem in Africa with a long-term impact on the availability of trained clinicians, neurologists and movement disorders experts and hence the diagnosis and provision of care for persons with PD on the continent. According to the WHO in 2017, there are only 0.03 neurologists per 100,000 population in Africa which is much less than what obtains in Southeast Asia and Europe (0.07 and 4.84 per 100,000 population respectively)^9^. In a recent comprehensive web-based survey conducted across 28 countries in Africa, the reported number of neurologists per million population ranged from 0.05 (Malawi) to 43.97 (Egypt)^20, 21, 199, 200^.

Providing evidence-based treatment and improved care options for patients is driven by research. Our review showed that data on patient-centred care is lacking. The availability and accessibility of PD therapy are still a major challenge in Africa. Looking for local and more accessible alternatives to levodopa such as mucuna pruriens has been explored^206^. Multi-sectoral engagement involving governments, international foundations, and pharmaceutical industries will be required to bridge this gap akin to strategies used to provide anti-retroviral therapy (ARV) for Africa.

In addition, data on patient engagement and QoL are lacking. This is probably due to stigmatization linked to the disease resulting in reduced interaction with the healthcare service. Furthermore, the lack of validated assessment tools which may not be available in the local language of the patient makes data collection difficult. This is a huge problem in Africa as there are so many languages being spoken across the different regions of the continent. Resolving these issues will require contacting stakeholders (persons living with PD, patient advocates, academic neurologists), international organisations and policy makers to conduct research targeted at the provision of validated tools in the more common African languages and the provision of affordable treatment^20^.

Researchable gaps from this review include patient-centred care and outcomes including quality of life, patient’s knowledge, and lived experiences. Other gaps include up-to-date epidemiology, education and training of healthcare professionals, identifying risk factors/biomarkers for PD and management and access to care for people with PD. To fill these gaps, research fragmentation which is a major problem across the continent should be addressed. This can be resolved by developing interconnectivity between centres and leveraging on efforts ongoing within the continent for example deployment of clinical registries, virtual collaborative centres and patients and public engagement. Also, regional integration is required in the areas of education and training, research, advocacy as well as health policies. This can be strengthened by the support of international collaborations with international organizations and building connections with LMICs in other parts of the world, who may have similar challenges in the short term.

Our recommendations are as follows: upgrade of healthcare and research systems within Africa. These include better training for allied healthcare practitioners in the area of early recognition and referral of persons with PD, increased training of physicians and neurologists as well as planned retention of these professionals within the continent. In addition, improved access to affordable medications is an unmet need in the care of persons with PD in the continent and this should be the top priority. Improvements in patient education, awareness and recognition of symptoms, early diagnosis and participation in research can only be sustained when persons with PD have access to available treatment. We also recommend intra-continental and international collaboration as this is key to bridging deficiencies in research output in low- and low-middle-income countries (LMICs) as it enables the pooling of resources, expertise, and infrastructure to enhance and upgrade the research enterprise.

It is of note that global advancements in genetics and molecular biology continue to drive our understanding of complex diseases such as PD, and such knowledge is required for understanding PD in Africa, but this alone cannot overcome the challenges and obstacles to care faced by Africans living with PD. Africa requires improved infrastructure for more up-to-date epidemiologic and improved healthcare systems to bridge the current gaps in PD awareness, education, treatment and research.

### Strengths and limitations

A major strength of our review is that we have used eleven databases for our search and captured most of the included articles providing data. Another strength of our review is the collaboration between African countries. This is the largest collaborative effort to date, bringing together PD enthusiasts from all over Africa. Authors found studies from grey literature and local journals in their countries. This increased inclusion of studies and allowed us to identify work done in PD in the continent. Our study is limited by high-quality studies due to variations in methods which is probably a reflection of the complex and diverse nature of the continent.

### Conclusion

This review has shown that there is a growing body of research on PD in Africa, with the quality of the studies ranging from low to moderate. Despite this growth, research output from the continent is poor compared to that obtained from other parts of the world. All these are attributable to a lack of economic and political wealth, resources including personnel and infrastructure, and lack of policies targeting education and training and poor health systems including health financing.

## Data Availability

All data generated or analysed during this study are included in this published article (and its supplementary information files).

## References

1. GBD 2015 Neurological Disorders Collaborator Group. Global, regional, and national burden of neurological disorders during 1990–2015: a systematic analysis for the Global Burden of Disease Study 2015. The Lancet Neurology 16, 877-897 (2017).

2. GBD 2016 Parkinson’s disease Collaborators. Global, regional, and national burden of Parkinson’s disease, 1990–2016: a systematic analysis for the Global Burden of Disease Study 2016. The Lancet Neurology 17, 939-953 (2018).

3. The Lancet, N. Parkinson’s disease needs an urgent public health response. Lancet Neurol 21, 759 (2022).

4. WHO. Parkinson disease: a public health approach. Technical brief. Geneva: World Health Organization; 2022. (2022).

5. Twelves, D., Perkins, K. S. M., Counsell, C. Systematic review of incidence studies of Parkinson’s disease. Mov Disord 18, 19–31 (2003).

6. von Campenhausen, S., et al. Prevalence and incidence of Parkinson’s disease in Europe. Eur Neuropsychopharmacol 15, 473–490 (2005).

7. Dorsey, E. R., et al. Projected number of people with Parkinson disease in the most populous nations, 2005 through 2030. Neurology 68, 384-386 (2007).

8. GBD 2016 Collaborators. Global, regional, and national burden of neurological disorders, 1990–2016: a systematic analysis for the Global Burden of Disease Study 2016. The Lancet Neurology 18, 459-480 (2019).

9. World Health Organisation. Atlas: Country resources for neurological disorders - 2nd ed. Geneva: World Health Organisation Licence: CC BY-NC-SA 3.0 IGO, (2017).

10. Nations, U. Data Portal. Population Division (2022).

11. Pringsheim, T., Jette, N., Frolkis, A., Steeves, T. D. The prevalence of Parkinson’s disease: a systematic review and meta-analysis. Mov Disord 29, 1583–1590 (2014).

12. Ben-Joseph, A., Marshall, C. R., Lees, A. J., Noyce, A. J. Ethnic Variation in the Manifestation of Parkinson’s Disease: A Narrative Review. J Parkinsons Dis 10, 31–45 (2020).

13. Wright Willis, A., et al. Geographic and ethnic variation in Parkinson disease: a population-based study of US Medicare beneficiaries. Neuroepidemiology 34, 143–151 (2010).

14. Dotchin, C., et al. The prevalence of Parkinson’s disease in rural Tanzania. Mov Disord 23, 1567–1672 (2008).

15. Van Den Eeden, S. K., et al. Incidence of Parkinson’s disease: variation by age, gender, and race/ethnicity. Am J Epidemiol 157, 1015–1022 (2003).

16. Okubadejo, N. U., Bower, J. H., Rocca, W. A., Maraganore, D. M. Parkinson’s disease in Africa: A systematic review of epidemiologic and genetic studies. Mov Disord 21, 2150–2156 (2006).

17. Williams, U., Bandmann, O., Walker, R. Parkinson’s disease in Sub-Saharan Africa: A review of epidemiology, genetics and access to care. (2018).

18. Dotchin, C. L., Msuya, O., Walker, R. W. The challenge of Parkinson’s disease management in Africa. Age Ageing 36, 122–127 (2007).

19. Pearce, V., Wilson, I. Parkinson’s disease in Africa. Oxford University Press; 2007.

20. Hamid, E., et al. Availability of Therapies and Services for Parkinson’s Disease in Africa: A Continent-Wide Survey. Mov Disord 36, 2393–2407 (2021).

21. Hamid, E., et al. The Gaps and Prospects of Movement Disorders Education and Research in Africa: A Continental Survey. Mov Disord 38, 178–184 (2023).

22. LeWitt, P. A., Chaudhuri, K. R. Unmet needs in Parkinson disease: Motor and non-motor. Parkinsonism Relat Disord 80 Suppl 1, S7–S12 (2020).

23. Moher, D., Liberati, A., Tetzlaff, J., Altman, D. G. Preferred reporting items for systematic reviews and meta-analyses: the PRISMA statement. Ann Intern Med 151, 264–269, w264 (2009).

24. Wells, G., et al. The Newcastle–Ottawa Scale (NOS) for Assessing the Quality of Non-Randomized Studies in Meta-Analysis. ᅟ, (2000).

25. Gouider-Khouja, N., Belal, S., Hamida, M. B., Hentati, F. Clinical and genetic study of familial Parkinson’s disease in Tunisia. Neurology 54, 1603–1609 (2000).

26. Gouider-Khouja, N., et al. Autosomal recessive parkinsonism linked to parkin gene in a Tunisian family. Clinical, genetic and pathological study. Parkinsonism & Related Disorders 9, 247–251 (2003).

27. Atadzhanov, M., Zumla, A., Mwaba, P. Study of familial Parkinson’s disease in Russia, Uzbekistan, and Zambia. Postgrad Med J 81, 117–121 (2005).

28. Lesage, S., et al. G2019S LRRK2 mutation in French and North African families with Parkinson’s disease. Ann Neurol 58, 784–787 (2005).

29. Ishihara, L., et al. Clinical features of Parkinson disease patients with homozygous leucine-rich repeat kinase 2 G2019S mutations. Arch Neurol 63, 1250–1254 (2006).

30. Leutenegger, A.-l., et al. Juvenile-Onset Parkinsonism as a Result of the First Mutation in the Adenosine Triphosphate Orientation Domain of PINK1. Archives of neurology 63, 1257–1261 (2006).

31. Ishihara, L., et al. Screening for Lrrk2 G2019S and clinical comparison of Tunisian and North American Caucasian Parkinson’s disease families. Mov Disord 22, 55–61 (2007).

32. Hulihan, M. M., et al. LRRK2 Gly2019Ser penetrance in Arab–Berber patients from Tunisia: a case-control genetic study. The Lancet Neurology 7, 591–594 (2008).

33. Lesage, S., et al. Is the common LRRK2 G2019S mutation related to dyskinesias in North African Parkinson disease? Neurology 71, 1550–1552 (2008).

34. Okubadejo, N., et al. Analysis of Nigerians with apparently sporadic Parkinson disease for mutations in LRRK2, PRKN and ATXN3. PLoS One 3, e3421 (2008).

35. Warren, L., et al. A founding LRRK2 haplotype shared by Tunisian, US, European and Middle Eastern families with Parkinson’s disease. Parkinsonism Relat Disord 14, 77–80 (2008).

36. Bardien, S., et al. Molecular analysis of the parkin gene in South African patients diagnosed with Parkinson’s disease. Parkinsonism Relat Disord 15, 116–121 (2009).

37. Cazeneuve, C., et al. A new complex homozygous large rearrangement of the PINK1 gene in a Sudanese family with early onset Parkinson’s disease. Neurogenetics 10, 265–270 (2009).

38. Keyser, R. J., et al. Identification of a novel functional deletion variant in the 5’-UTR of the DJ-1 gene. BMC Med Genet 10, 105 (2009).

39. Nishioka, K., et al. A comparative study of LRRK2, PINK1 and genetically undefined familial Parkinson’s disease. J Neurol Neurosurg Psychiatry 81, 391-395 (2009).

40. Vilarino-Guell, C., et al. ATP13A2 variability in Parkinson disease. Hum Mutat 30, 406–410 (2009).

41. Bardien, S., et al. LRRK2 G2019S mutation: frequency and haplotype data in South African Parkinson’s disease patients. J Neural Transm (Vienna) 117, 847–853 (2010).

42. Belarbi, S., et al. LRRK2 G2019S mutation in Parkinson’s disease: a neuropsychological and neuropsychiatric study in a large Algerian cohort. Parkinsonism Relat Disord 16, 676–679 (2010).

43. Jasinska-Myga, B., et al. Comprehensive sequencing of the LRRK2 gene in patients with familial Parkinson’s disease from North Africa. Mov Disord 25, 2052–2058 (2010).

44. Keyser, R. J., et al. Assessing the prevalence of PINK1 genetic variants in South African patients diagnosed with early-and late-onset Parkinson’s disease. Biochem Biophys Res Commun 398, 125–129 (2010).

45. Keyser, R. J., et al. Analysis of exon dosage using MLPA in South African Parkinson’s disease patients. Neurogenetics 11, 305–312 (2010).

46. Lesage, S., et al. Parkinson’s disease-related LRRK2 G2019S mutation results from independent mutational events in humans. Hum Mol Genet 19, 1998–2004 (2010).

47. Nishioka, K., et al. Genetic variation of the mitochondrial complex I subunit NDUFV2 and Parkinson’s disease. Parkinsonism Relat Disord 16, 686–687 (2010).

48. Nishioka, K., et al. Glucocerebrosidase mutations are not a common risk factor for Parkinson disease in North Africa. Neurosci Lett 477, 57–60 (2010).

49. Troiano, A. R., et al. Low disease risk in relatives of north african lrrk2 Parkinson disease patients. Neurology 75, 1118–1119 (2010).

50. Ben Sassi, S., et al. Cognitive dysfunction in Tunisian LRRK2 associated Parkinson’s disease. Parkinsonism Relat Disord 18, 243–246 (2011).

51. Hashad, D. I., et al. G2019S mutation of the leucine-rich repeat kinase 2 gene in a cohort of Egyptian patients with Parkinson’s disease. Genet Test Mol Biomarkers 15, 861–866 (2011).

52. Keyser, R. J., Oppon, E., Carr, J. A., Bardien, S. Identification of Parkinson’s disease candidate genes using CAESAR and screening of MAPT and SNCAIP in South African Parkinson’s disease patients. J Neural Transm (Vienna) 118, 889–897 (2011).

53. Haylett, W. L., et al. Mutations in the parkin gene are a minor cause of Parkinson’s disease in the South African population. Parkinsonism Relat Disord 18, 89–92 (2012).

54. Yonova-Doing, E., et al. Analysis of LRRK2, SNCA, Parkin, PINK1, and DJ-1 in Zambian patients with Parkinson’s disease. Parkinsonism Relat Disord 18, 567-571 (2012).

55. Bardien, S., et al. Patient-control association study of the Leucine-Rich repeat kinase 2 (LRRK2) gene in South African Parkinson’s disease patients. Mov Disord 28, 2039–2040 (2013).

56. Trinh, J., et al. STX6 rs1411478 is not associated with increased risk of Parkinson’s disease. Parkinsonism Relat Disord 19, 563–565 (2013).

57. Blanckenberg, J., Ntsapi, C., Carr, J. A., Bardien, S. EIF4G1 R1205H and VPS35 D620N mutations are rare in Parkinson’s disease from South Africa. Neurobiol Aging 35, 445 e441–443 (2014).

58. Glanzmann, B., Lombard, D., Carr, J., Bardien, S. Screening of two indel polymorphisms in the 5’UTR of the DJ-1 gene in South African Parkinson’s disease patients. J Neural Transm (Vienna) 121, 135–138 (2014).

59. Hentati, F., et al. LRRK2 parkinsonism in Tunisia and Norway: a comparative analysis of disease penetrance. Neurology 83, 568–569 (2014).

60. Trinh, J., et al. A Comparative study of Parkinson’s disease and leucine-rich repeat kinase 2 p.G2019S parkinsonism. Neurobiol Aging 35, 1125-1131 (2014).

61. van der Merwe, C., et al. Mitochondrial impairment observed in fibroblasts from South African Parkinson’s disease patients with parkin mutations. Biochem Biophys Res Commun 447, 334–340 (2014).

62. El Desoky, E., Khedr, E., Khalil, M., Gasser, T. Genetic Analysis of Leucin-Rich Repeat Kinase 2 (LRRK2) G2019S Mutation in a Sample of Egyptian Patients with Parkinson’s Disease, a Pilot Study. British Journal of Medicine and Medical Research 5, 404–408 (2015).

63. Ben El Haj, R., et al. A Novel Homozygous p.L539F Mutation Identified in PINK1 Gene in a Moroccan Patient with Parkinsonism. Biomed Res Int 2016, 3460234 (2016).

64. Sayad, M., et al. Greater improvement in LRRK2 G2019S patients undergoing Subthalamic Nucleus Deep Brain Stimulation compared to non-mutation carriers. BMC Neurosci 17, 6 (2016).

65. van der Merwe, C., Carr, J., Glanzmann, B., Bardien, S. Exonic rearrangements in the known Parkinson’s disease-causing genes are a rare cause of the disease in South African patients. Neurosci Lett 619, 168–171 (2016).

66. Barkhuizen, M., Anderson, D. G., van der Westhuizen, F. H., Grobler, A. F. A molecular analysis of the GBA gene in Caucasian South Africans with Parkinson’s disease. Mol Genet Genomic Med 5, 147–156 (2017).

67. Ben El Haj, R., et al. Evidence for prehistoric origins of the G2019S mutation in the North African Berber population. PLoS One 12, e0181335 (2017).

68. Bouhouche, A., et al. LRRK2 G2019S Mutation: Prevalence and Clinical Features in Moroccans with Parkinson’s Disease. Parkinsons Dis 2017, 2412486 (2017).

69. Landoulsi, Z., et al. Using KASP technique to screen LRRK2 G2019S mutation in a large Tunisian cohort. BMC Med Genet 18, 70 (2017).

70. Ben Romdhan, S., et al. LRRK2 G2019S Parkinson’s disease with more benign phenotype than idiopathic. Acta Neurol Scand 138, 425–431 (2018).

71. Fahmy, E., et al. Interleukin-18 promoter polymorphisms and idiopathic Parkinson disease: an Egyptian study. Acta Neurol Belg 119, 219–224 (2018).

72. Kuipers, D. J. S., et al. PTRHD1 Loss-of-function mutation in an african family with juvenile-onset Parkinsonism and intellectual disability. Mov Disord 33, 1814–1819 (2018).

73. Okubadejo, N. U., et al. Leucine rich repeat kinase 2 (LRRK2) GLY2019SER mutation is absent in a second cohort of Nigerian Africans with Parkinson disease. PLoS One 13, e0207984 (2018).

74. Ibrahim A Etedal Ahmed Abdalaziz, A. S. Pink1 type of Early Onset Parkinson’s disease(EOPD)in Sudanese patients, 2018. Sudan Journal of Medical Sciences (2019).

75. du Toit, N., et al. Frequency of the LRRK2 G2019S mutation in South African patients with Parkinson’s disease. Neurogenetics 20, 215–218 (2019).

76. Mahungu, A. C., et al. Screening of the glucocerebrosidase (GBA) gene in South Africans of African ancestry with Parkinson’s disease. Neurobiol Aging 88, 156 e111–156 e114 (2019).

77. Dekker, M. C. J., et al. PRKN-related familial Parkinson’s disease: First molecular confirmation from East Africa. Parkinsonism Relat Disord 73, 14–15 (2020).

78. du Plessis, S., et al. Association Between a Variable Number Tandem Repeat Polymorphism Within the DAT1 Gene and the Mesolimbic Pathway in Parkinson’s Disease. Front Neurol 11, 982 (2020).

79. Luth, T., et al. Age at Onset of LRRK2 p.Gly2019Ser Is Related to Environmental and Lifestyle Factors. Mov Disord 35, 1854-1858 (2020).

80. Oluwole, O. G., et al. Targeted next-generation sequencing identifies novel variants in candidate genes for Parkinson’s disease in Black South African and Nigerian patients. BMC Med Genet 21, 23 (2020).

81. Rebai, A., et al. Effects of glutathione S-transferase M1 and T1 deletions on Parkinson’s disease risk among a North African population. Rev Neurol (Paris) (2020).

82. El-Tallawy, H. N., et al. Study of cognitive impairment and genetic polymorphism of SLC41A1 (rs11240569 allele) in Parkinson’s disease in Upper Egypt: case-control study. The Egyptian Journal of Neurology, Psychiatry and Neurosurgery 57, (2021).

83. Milanowski, L. M., et al. Early-Onset Parkinson Disease Screening in Patients From Nigeria. Front Neurol 11, 594927 (2021).

84. Rizig, M., et al. Negative screening for 12 rare LRRK2 pathogenic variants in a cohort of Nigerians with Parkinson’s disease. Neurobiol Aging 99, 101 e115–101 e119 (2021).

85. Sebate, B., et al. Prioritization of candidate genes for a South African family with Parkinson’s disease using in-silico tools. PLoS One 16, e0249324 (2021).

86. Smaili, I., et al. Gene Panel Sequencing Identifies Novel Pathogenic Mutations in Moroccan Patients with Familial Parkinson Disease. J Mol Neurosci 71, 142–152 (2021).

87. Okubadejo, N. U., et al. APOE E4 is associated with impaired self-declared cognition but not disease risk or age of onset in Nigerians with Parkinson’s disease. NPJ Parkinsons Dis 8, 155 (2022).

88. Nasri, A., et al. Heart rate variability and sympathetic skin response for the assessment of autonomic dysfunction in leucine-rich repeat kinase 2 associated Parkinson’s disease. Neurophysiol Clin 52, 81–93 (2022).

89. Muller-Nedebock, A. C., et al. Increased blood-derived mitochondrial DNA copy number in African ancestry individuals with Parkinson’s disease. Parkinsonism Relat Disord 101, 1–5 (2022).

90. Bakhit, Y., et al. Methylation of alpha-synuclein in a Sudanese cohort. Parkinsonism Relat Disord 101, 6–8 (2022).

91. Ait Wahmane, S., et al. Parkinson’s disease: The prevalence of the LRRK2-G2019S mutation among the population of the South-West of Morocco. Gene Reports 27, (2022).

92. Okubadejo, N. U., Danesi, M. A. Frequency and predictors of autonomic dysfunction in Parkinson’s disease: a study of African patients in Lagos, Nigeria. Niger Postgrad Med J 11, 45–49 (2004).

93. Ishihara, L., et al. Clinical features of Parkinson disease patients with homozygous leucine-rich repeat kinase 2 G2019S mutations. Archives of neurology 63, 1250–1254 (2006).

94. Akinyemi, R. O., et al. Cognitive dysfunction in Nigerians with Parkinson’s disease. Mov Disord 23, 1378–1383 (2008).

95. Matuja, W. B., Aris, E. A. Motor and non-motor features of Parkinson’s disease. East Afr Med J 85, 3–9 (2008).

96. Atadzhanov, M., Mwaba, P. Disorders of the basal ganglia at the University Teaching Hospital, Lusaka. Medical Journal of Zambia 36, (2009).

97. Dotchin, C. L., Jusabani, A., Walker, R. W. Non-motor symptoms in a prevalent population with Parkinson’s disease in Tanzania. Parkinsonism Relat Disord 15, 457–460 (2009).

98. Khalid, K., et al. Clinical presentation of Parkinson’s disease among Sudanese patients. 4, (2009).

99. Okubadejo, N. U., Ojo, O. O., Oshinaike, O. O. Clinical profile of parkinsonism and Parkinson’s disease in Lagos, Southwestern Nigeria. BMC Neurol 10, 1 (2010).

100. Ojo, O. O., et al. Plasma homocysteine level and its relationship to clinical profile in Parkinson’s disease patients at the Lagos University Teaching Hospital. West Afr J Med 30, (2011).

101. Femi, O. L., Ibrahim, A., Aliyu, S. Clinical profile of parkinsonian disorders in the tropics: Experience at Kano, northwestern Nigeria. J Neurosci Rural Pract 3, 237–241 (2012).

102. Miller, N., et al. Assessment of speech in neurological disorders: Development of a Swahili screening test. South African Journal of Communication Disorders 59, (2012).

103. Moustapha, S. M., et al. Pneumophonic coordination impairments in parkinsonian dysarthria: importance of aerodynamic parameters measurements. West Afr J Med 31, 129–134 (2012).

104. Ojo, O. O., Okubadejo, N. U., Ojini, F. I., Danesi, M. A. Frequency of cognitive impairment and depression in Parkinson’s disease: A preliminary case-control study. Niger Med J 53, 65–70 (2012).

105. Aris, E., Dotchin, C. L., Gray, W. K., Walker, R. W. Autonomic function in a prevalent Tanzanian population with Parkinson’s disease and its relationship to disease duration and 5-year mortality. BMC research notes 6, 1–5 (2013).

106. Darwish, M. H., El-Tamawy, M. S., Ahmed, S. M., Rasmy, H. Somatosensory and Motor Systems Affect Postural Stability in Parkinson’s Disease Patients. Egyptian Journal of Neurology, Psychiatry & Neurosurgery 50, (2013).

107. Ojagbemi, A. Relationship between cognitive dysfunction and behavioural symptoms in Nigerian patients with Parkinson’s disease no dementia. J Parkinsons Dis 3, 293–300 (2013).

108. Ojagbemi, A. A., Akinyemi, R. O., Baiyewu, O. Neuropsychiatric symptoms in Nigerian patients with Parkinson’s disease. Acta Neurol Scand 128, 9–16 (2013).

109. Okunoye, O. C., Asekomeh, G. s. E. Depression among patients with Parkinson’s disease in a Nigerian tertiary hospital. Nigerian Health Journal 13, 96–103 (2013).

110. Atadzhanov, M. Phenotypic Characteristics of Zambian patients with Parkinson’s Disease. Medical Journal of Zambia 41, 162–167 (2014).

111. Cilia, R., et al. The modern pre-levodopa era of Parkinson’s disease: insights into motor complications from sub-Saharan Africa. Brain 137, 2731–2742 (2014).

112. Cubo, E., et al. Comparison of the clinical profile of Parkinson’s disease between Spanish and Cameroonian cohorts. J Neurol Sci 336, 122–126 (2014).

113. Gaida, R., Truter, I. Preliminary Investigation of Risk Factors Causing Dyskinesias in Parkinson’s Disease in South Africa. Tropical Journal of Pharmaceutical Research 13, (2014).

114. Lf, O. Gastrointestinal complications in newly diagnosed Parkinson’s disease: A case-control study. Tropical Gastroenterology 35, 227–231 (2014).

115. Okunoye, O. C. Non-motor features in Parkinson’s disease patients attending neurology clinic at a tertiary institution in Nigeria: a preliminary report. The Nigerian Health Journal 14, 114 (2014).

116. Regragui, W., et al. Profile of idiopathic parkinson’s disease in Moroccan patients. Int Arch Med 7, 10 (2014).

117. Kisoli, A., et al. Levels of functional disability in elderly people in Tanzania with dementia, stroke and Parkinson’s disease. Acta Neuropsychiatr 27, 206–212 (2015).

118. Maiga, B., et al. Sleep quality assessment in 35 Parkinson’s disease patients in the Fann Teaching Hospital, Dakar, Senegal. Rev Neurol (Paris>) 172, 242–247 (2015).

119. Smith, M., Modi, G. The clinical profile of idiopathic Parkinson’s disease in a South African hospital complex-the influence of ethnicity and gender. African Journal of Neurological Sciences 35, 73–79 (2015).

120. Farombi, T. H., Owolabi, M. O., Ogunniyi, A. Falls and Their Associated Risks in Parkinson’s Disease Patients in Nigeria. J Mov Disord 9, 160–165 (2016).

121. Owolabi, L. F., Nagoda, M., Babashani, M. Pulmonary function tests in patients with Parkinson’s disease: A case-control study. Niger J Clin Pract 19, 66–70 (2016).

122. Assadeck, H., et al. Clinical Profile of Parkinson’s Disease: Experience of Niger. J Neurosci Rural Pract 9, 214–218 (2018).

123. Shalash, A. S., et al. Non-Motor Symptoms as Predictors of Quality of Life in Egyptian Patients With Parkinson’s Disease: A Cross-Sectional Study Using a Culturally Adapted 39-Item Parkinson’s Disease Questionnaire. Front Neurol 9, 357 (2018).

124. Amod, F. H., Bhigjee, A. I. Clinical series of Parkinson’s disease in KwaZulu-Natal, South Africa: Retrospective chart review. J Neurol Sci 401, 62-65 (2019).

125. Arabambi, B., Oshinaike, O., Ogun, S. A. Profile of Nonmotor Symptoms and the Association with the Quality of Life of Parkinson’s Disease Patients in Nigeria. Niger Med J 60, 273–278 (2019).

126. El Otmani, H., et al. Impulse control disorders in Parkinson disease: A cross-sectional study in Morocco. Rev Neurol (Paris) 175, 233–237 (2019).

127. Hirsi, J. O., Yifru, Y. M., Metaferia, G. Z., Bower, J. H. Prevalence of pain in patients with Parkinson’s disease in Addis Ababa, Ethiopia. Parkinsonism Relat Disord 61, 214–218 (2019).

128. Melka, D., Tafesse, A., Bower, J. H., Assefa, D. Prevalence of sleep disorders in Parkinson’s disease patients in two neurology referral hospitals in Ethiopia. BMC Neurol 19, 205 (2019).

129. Oparah, S., Mwankon, J. P. Clinical Profile of Parkinson’s Disease in Calabar, Southern Nigeria. Journal of Research in Basic and Clinical Sciences 1, 80–84 (2019).

130. Rasaholiarison, N. F., Razafimahefa, J., Rakotomanana, J. L., Tehindrazanarivelo, A. D. Frequency and clinical profile of Parkinson’s disease and other Parkinsonian syndromes seen in the Department of Neurology at the Befelatanana Hospital Antananarivo. Pan Afr Med J 33, 2019 (2019).

131. Achbani, A., et al. Gender and Age Difference in Clinical Features and severity of Parkinson’s Disease: A Cross-Sectional Study in Southern Morocco. Archives of Neuroscience 7, (2020).

132. Cilia, R., et al. Natural history of motor symptoms in Parkinson’s disease and the long-duration response to levodopa. Brain (London, England: 1878) 143, 2490-2501 (2020).

133. Melka, D., Tafesse, A., Sheferaw, S. Prevalence and determinants of fatigue among Parkinsn’s disease patients in Ethiopia. Ethipian Med J 58, 125–131 (2020).

134. Nelson, G., et al. Validation of Parkinson’s Disease-Related Questionnaires in South Africa. Parkinson’s disease 2020, 1–9 (2020).

135. Ojo, O. O., et al. The Nigeria Parkinson Disease Registry: Process, Profile, and Prospects of a Collaborative Project. Mov Disord 35, 1315–1322 (2020).

136. Shalash, A., et al. Translation, Validation, Diagnostic Accuracy, and Reliability of Screening Questionnaire for Parkinsonism in Three African Countries. Journal of Parkinson’s disease 10, 1113–1122 (2020).

137. Ahmadou, T. M., et al. Study of Unilateral Spatial Neglect in Parkinson’s Patients. Acta Neuropsychologica 19, 219–229 (2021).

138. Amod, F. H., Bhigjee, A. I., Moodley, A. Does antiretroviral therapy alter the course of Parkinson’s disease in people living with HIV? J Neurovirol 27, 595–600 (2021).

139. Ayele, B. A., et al. Non-Motor Symptoms and Associated Factors in Parkinson’s Disease Patients in Addis Ababa, Ethiopia: A Multicenter Cross-Sectional Study. Ethiop J Health Sci 31, 837-846 (2021).

140. Dlamini, W. W., et al. A Rapid Motor Task-Based Screening Tool for Parkinsonism in Community-Based Studies. Frontiers in neurology 12, 653066–653066 (2021).

141. El Otmani, H., et al. No impact of confinement during COVID-19 pandemic on anxiety and depression in Parkinsonian patients. Revue neurologique 177, 272–274 (2021).

142. Elsherif, M., et al. Prevalence of abnormal pulmonary functions in Parkinson’s disease and its correlation to the disease severity and quality of life. Medical Science 25, 1739–1747 (2021).

143. Fothergill-Misbah, N., et al. “Old people problems”, uncertainty and legitimacy: Challenges with diagnosing Parkinson’s disease in Kenya. Social science & medicine (1982) 282, 114148–114148 (2021).

144. Nasri, A., et al. Atypical parkinsonian syndromes in a North African tertiary referral center. Brain and behavior 11, e01924-n/a (2021).

145. Ojo, O. O., et al. A Cross-Sectional Comprehensive Assessment of the Profile and Burden of Non-motor Symptoms in Relation to Motor Phenotype in the Nigeria Parkinson Disease Registry Cohort. Movement disorders clinical practice (Hoboken, NJ) 8, 1206–1215 (2021).

146. Shalash, A., et al. A 6-month longitudinal study on worsening of Parkinson’s disease during the COVID-19 pandemic. NPJ Parkinsons Dis 8, 111 (2022).

147. Talaat, M. A. M., Elfatatry, A., Noor, N. M. I. M., Eldeeb, M. Assessment of postural instability in Parkinson’s disease patients. The Egyptian Journal of Otolaryngology 37, (2021).

148. Helmy, A., et al. Baseline predictors of progression of Parkinson’s disease in a sample of Egyptian patients: clinical and biochemical. Egypt J Neurol Psychiatr Neurosurg 58, 9 (2022).

149. Emad, E. M., Elmotaym, A. S. E., Ghonemy, M. m. A., Badawy, A. E. The effect of hypocalcemia on motor symptoms of Parkinson’s disease. The Egyptian Journal of Neurology, Psychiatry and Neurosurgery 58, (2022).

150. Ayele, B. A., Tesfaye, H., Wuhib, Z. M., Zenebe, G. Factors Associated with EEG Slowing in Individuals with Parkinson’s Disease. Ethiop J Health Sci 32, 73–80 (2022).

151. Mengesha, A. T. Frequency and Factors Associated with Orthostatic Hypotension in Individuals with Parkinson’s Disease: A Case-Control Observational Study. Ethiop J Health Sci 32, 1167–1174 (2022).

152. El-Kattan, M. M., et al. Optical coherence tomography in patients with Parkinson’s disease. The Egyptian Journal of Neurology, Psychiatry and Neurosurgery 58, (2022).

153. El Mokadem, M. O., Hassan, A., Hussein, M., Mohamed, Y. M. The potential role of 2D-speckle tracking echocardiography for detecting left ventricular systolic dysfunction in patients with Parkinson’s disease: a case control study. Acta Cardiol 76, 979–986 (2021).

154. Shaheen, S., et al. Screening for non-motor symptoms in Egyptian patients with Parkinson’s disease. The Egyptian Journal of Neurology, Psychiatry and Neurosurgery 58, (2022).

155. Elshamy, A. M., Mohamed, E. S., Al-Malt, A. M., Ragab, O. A. Sexual Dysfunction Among Egyptian Idiopathic Parkinson’s Disease Patients. J Geriatr Psychiatry Neurol 35, 816–822 (2022).

156. Shalash, A. S., et al. Non-motor symptoms in essential tremor, akinetic rigid and tremor-dominant subtypes of Parkinson’s disease. PLoS One 16, e0245918 (2021).

157. Ashok, P. P., Radhakrishnan, K., Sridharan, R., Mousa, M. E. Epidemiology of Parkinson’s disease in Benghazi, North-East Libya. Clin Neurol Neurosurg 88, 109–113 (1986).

158. Schoenberg, B. S., et al. Comparison of the prevalence of Parkinson’s disease in black populations in the rural United States and in rural Nigeria: door-to-door community studies. Neurology 38, 645–646 (1988).

159. Tekle-Haimanot, R., et al. Community-based study of neurological disorders in rural central Ethiopia. Neuroepidemiology 9, 263–277 (1990).

160. Attia Romdhane, N., et al. Prevalence study of neurologic disorders in Kelibia (Tunisia). Neuroepidemiology 12, 285–299 (1993).

161. Kengne, A. P., Dzudie, A., Dongmo, L. Epidemiological features of degenerative brain diseases as they occurred in Yaounde referral hospitals over a 9-year period. Neuroepidemiology 27, 208–211 (2006).

162. Ekenze, O. S., Onwuekwe, I. O., Ezeala Adikaibe, B. A. Profile of neurological admissions at the University of Nigeria Teaching Hospital Enugu. Niger J Med 19, 419–422 (2010).

163. Dewhurst, F., et al. The prevalence of neurological disorders in older people in Tanzania. Acta Neurol Scand 127, 198–207 (2013).

164. El-Tallawy, H. N., et al. Door-to-door survey of major neurological disorders (project) in Al Quseir City, Red Sea Governorate, Egypt. Neuropsychiatr Dis Treat 9, 767–771 (2013).

165. El-Tallawy, H. N., et al. Prevalence of Parkinson’s disease and other types of Parkinsonism in Al Kharga district, Egypt. Neuropsychiatr Dis Treat 9, 1821–1826 (2013).

166. Callixte, K. T., et al. The pattern of neurological diseases in elderly people in outpatient consultations in Sub-Saharan Africa. BMC Res Notes 8, 159 (2015).

167. Khedr, E. M., et al. Prevalence of Parkinsonism and Parkinson’s disease in Qena governorate/Egypt: a cross-sectional community-based survey. Neurol Res 37, 607–618 (2015).

168. Cubo, E., et al. The Burden of Movement Disorders in Cameroon: A Rural and Urban-Based Inpatient/Outpatient Study. Mov Disord Clin Pract 4, 568–573 (2017).

169. Otubogun, F. M., Akinyemi, R., Ogunniyi, S. Burden of adult neurological diseases in Odeda Area, Southwest Nigeria. BMJ neurology open 2, e000062–e000062 (2020).

170. Sejvar, J., et al. Neurologic manifestations associated with an outbreak of typhoid fever, Malawi--Mozambique, 2009: an epidemiologic investigation. PLoS One 7, e46099 (2012).

171. van der Merwe, C., et al. Factors influencing the development of early-or late-onset Parkinson’s disease in a cohort of South African patients. S Afr Med J 102, 848–851 (2012).

172. Ogunrin, O., et al. Trace Metals in Patients with Parkinson’s Disease: A Multi-Center Case-Control Study of Nigerian Patients. Journal of Neurology and Epidemiology 1, 31–38 (2013).

173. Ashour, S., et al. A Study of Extrapyramidal Manifestations Accompanying Decompensated Viral Hepatic Cirrhosis Patients. Rev Recent Clin Trials 12, 162–167 (2017).

174. Rosler, T. W., et al. K-variant BCHE and pesticide exposure: Gene-environment interactions in a case-control study of Parkinson’s disease in Egypt. Sci Rep 8, 16525 (2018).

175. Dlamini, W. W., Nelson, G., Nielsen, S. S., Racette, B. A. Manganese exposure, parkinsonian signs, and quality of life in South African mine workers. Am J Ind Med 63, 36–43 (2020).

176. Kissini, N., et al. Parkinsonism and chronic manganese exposure: Pilot study with clinical, environmental and experimental evidence. Clinical Parkinsonism & Related Disorders 3, (2020).

177. Rebai, A., et al. GC-MS Based Metabolic Profiling of Parkinson’s Disease with Glutathione S-transferase M1 and T1 Polymorphism in Tunisian Patients. Comb Chem High Throughput Screen 23, 1041–1048 (2020).

178. Racette, B. A., et al. Severity of parkinsonism associated with environmental manganese exposure. Environ Health 20, 27 (2021).

179. Ghit, A., Deeb, H. E. Cytokines, miRNAs, and Antioxidants as Combined Non-invasive Biomarkers for Parkinson’s Disease. J Mol Neurosci 72, 1133–1140 (2022).

180. Khedr, E. M., et al. Gut microbiota in Parkinson’s disease patients: hospital-based study. The Egyptian Journal of Neurology, Psychiatry and Neurosurgery 57, (2021).

181. El-Kattan, M. M., Rashed, L. A., Shazly, S. R., Ismail, R. S. Relation of serum level of tumor necrosis factor-alpha to cognitive functions in patients with Parkinson’s disease. The Egyptian Journal of Neurology, Psychiatry and Neurosurgery 58, (2022).

182. Achbani, A., et al. Risk Factors of Parkinson’s Disease: A Case-Control Study in Moroccan Patients. Archives of Neuroscience 9, (2022).

183. Mshana, G., Dotchin, C. L., Walker, R. W. ’We call it the shaking illness’: perceptions and experiences of Parkinson’s disease in rural northern Tanzania. BMC Public Health 11, 219 (2011).

184. Okunoye, O. C., et al. Profile of Generic and Disease-Specific Health-Related Quality of Life Among Nigerians with Parkinsons Disease. The Nigerian Health Journal 14, 79–86 (2014).

185. Okunoye, O. C., Asekomeh, G. s. E., Owolabi, M., Otike-Odibi, B. I. Determinants of health-related quality of life among Nigerian-Africans with Parkinson’s disease. Port Harcourt Medical Journal 8, 50–60 (2014).

186. Kaddumukasa, M., et al. Knowledge and Attitudes of Parkinson’s Disease in Rural and Urban Mukono District, Uganda: A Cross-Sectional, Community-Based Study. Parkinsons Dis 2015, 196150 (2015).

187. Mokaya, J., Gray, W. K., Carr, J. Beliefs, knowledge and attitudes towards Parkinson’s disease among a Xhosa speaking black population in South Africa: A cross-sectional study. Parkinsonism Relat Disord 41, 51–57 (2017).

188. Walga, T. K. Understanding the Experience and Perspectives of Parkinson’s Disease Patients’ Caregivers. Rehabil Res Pract 2019, 3082325 (2019).

189. El Semary, M., et al. Efficacy of Cognitive Rehabilitation on Functional Outcomes & Quality of Life in Parkinson’s Patients. International Journal of Pharmaceutical Research 12, (2020).

190. Shalash, A., et al. Mental Health, Physical Activity, and Quality of Life in Parkinson’s Disease During COVID-19 Pandemic. Movement disorders 35, 1097–1099 (2020).

191. Fothergill-Misbah, N., et al. The role of support groups in the management of Parkinson’s disease in Kenya: Sociality, information and legitimacy. Glob Public Health 1–11 (2021).

192. Shalash, A., et al. Sexual dysfunction in male patients with Parkinson’s disease: related factors and impact on quality of life. Neurol Sci 41, 2201–2206 (2020).

193. Dotchin, C., Jusabani, A., Walker, R. Three year follow up of levodopa plus carbidopa treatment in a prevalent cohort of patients with Parkinson’s disease in Hai, Tanzania. J Neurol 258, 1649–1656 (2011).

194. Mokaya, J., et al. The Accessibility of Parkinson’s Disease Medication in Kenya: Results of a National Survey. Mov Disord Clin Pract 3, 376–381 (2016).

195. Okubadejo, N. U., et al. A Nationwide Survey of Parkinson’s Disease Medicines Availability and Affordability in Nigeria. Mov Disord Clin Pract 6, 27–33 (2019).

196. Chahra, C., et al. The effect of Origanum majorana tea on motor and non-motor symptoms in patients with idiopathic Parkinson’s disease: A randomized controlled pilot study. Parkinsonism Relat Disord 91, 23–27 (2021).

197. Barichella, M., et al. Nutritional status and dietary habits in Parkinson’s disease patients in Ghana. Nutrition 29, 470–473 (2013).

198. Naeije, G., et al. Yield of training exchanges between Europe and Sub-Saharan Africa. Acta Neurol Belg 113, 31–34 (2013).

199. Cubo, E., et al. A Parkinson’s disease tele-education program for health care providers in Cameroon. J Neurol Sci 357, 285–287 (2015).

200. Cubo, E., et al. Telemedicine Enables Broader Access to Movement Disorders Curricula for Medical Students. Tremor Other Hyperkinet Mov (N Y) 7, 501 (2017).

201. Norman, B. P., et al. Early Onset Parkinson’s Disease in a family of Moroccan origin caused by a p.A217D mutation in PINK1: a case report. BMC Neurol 17, 153 (2017).

202. Odeniyi, O., Ojo, O., Odeniyi, I., Okubadejo, N. Association of serum uric acid and non-motor symptoms in Parkinson’s disease: A cross-sectional study from a movement disorders clinic in Lagos, Nigeria. Journal of Clinical Sciences 19, 104–109 (2022).

203. Sakadi, F., et al. Gait disorders in Parkinson’s disease: 19 observations at the University Hospital Center of Conakry. Movement disorders 33, (2018).

204. Lesage, S., et al. LRRK2 G2019S as a cause of Parkinson’s disease in North African Arabs. N Engl J Med 354, 422-423 (2006).

205. Lesage, S., et al. LRRK2 haplotype analyses in European and North African families with Parkinson disease: a common founder for the G2019S mutation dating from the 13th century. Am J Hum Genet 77, 330–332 (2005).

