## Supplemental Table 1 for "The State of Play of Parkinson’s Disease in Africa: A Systematic Review and Point of View"

Supplementary Table 1: Summary of included studies

| Study ID | Title and year | Theme | Design of study | African country(ies) defined by PD Cohort | Collaborating country | First Author’s name | Journal and impact factor  2022 | Source of funding |
| --- | --- | --- | --- | --- | --- | --- | --- | --- |
| 1 | Clinical and genetic study of familial Parkinson's disease in Tunisia (2000). | Genetics | Prospective  (Case- series)  PD=88 (21 families) | Tunisia | None | Gouider-Khouja et al | Neurology  9.9 | Research fellowship funding provided by the Restless legs Syndrome Foundation to W.B.J. M.A. is a NIH visiting fellow from Erciyes University in Turkey and partly supported by the Turkish Scientific and Technological Council. |
| 2 | Autosomal recessive parkinsonism linked to parkin gene in a Tunisian family. Clinical, genetic and pathological study (2003). | Genetics | Prospective  (Case of one family)  PD=3 siblings | Tunisia | Japan | Gouider-Khouja et al | Parkinsonism  and related  disorders  4.8 | Not reported |
| 3 | G2019S LRRK2 mutation in French and North African families with Parkinson's disease (2005). | Genetics | Prospective  (Familial)  PD=17 were African | Algeria Tunisia Morocco | Portugal  France | Lesage et al | Annals of Neurology  10.4 | European Grant APOPIS(European Union-Contract No. LSHM-CT-2003-503330 and Institut National de la Sante et de la Recherche Me´dicale/AFM Cohortes et Collections 2001 |
| 4 | Study of familial Parkinson's disease in Russia, Uzbekistan, and Zambia (2005) | Genetics | Prospective  Observational/  Descriptive  PD=52 | Zambia | Russia Uzbekistan | Atadzhanov et al | Postgraduate medical Journal  2.07 | Not reported |

Table 1(contd): Summary of included studies

| Study ID | Title and year | Theme | Design of study | African country(ies) defined by PD Cohort | Collaborating countries | First Author’s name | Journal and impact factor  2022 | Source of funding |
| --- | --- | --- | --- | --- | --- | --- | --- | --- |
| 5 | Juvenile-onset Parkinsonism as a result of the first mutation in the adenosine triphosphate orientation domain of PINK1(2006). | Genetics | Prospective  Case-control  PD=8 | Sudan | France Saudi Arabia | Leutenegger et al | Archives of Neurology  (now JAMA)  29.9 | Grant NS41723-01A1 from the National Institutes of Health. |
| 6 | Clinical features of Parkinson disease patients with homozygous leucine-rich repeat kinase 2 G2019S mutations (2006) | Genetics | Prospective  PD=26 | Morocco Tunisia Algeria | France, USA  Japan, UK  Algeria | Ishihara et al | Archives of Neurology  (now JAMA)  29.9 | Not reported |
| 7 | Screening for LRRK2 G2019S and clinical comparison of Tunisian and North American Caucasian Parkinson's disease families (2007) | Genetics | Prospective  Case-control  PD=197 | Tunisia | USA  Japan | Ishihara et al | Movement Disorders  9.6 | Not reported |
| 8 | LRRK2 Gly2019Ser penetrance in Arab-Berber patients from Tunisia: a case-control genetic study (2008). | Genetics | Case-control  PD=238 | Tunisia | UK  USA | Hulihan et al | Lancet Neurology  44.1 | GlaxoSmithKline, Institute of Neurology Tunis and Mayo Foundation |

Table 1(contd): Summary of included studies

| Study ID | Title and year | Theme | Design of study | African country(ies) defined by PD Cohort | Collaborating country | First Author’s name | Journal and impact factor  2022 | Source of funding |
| --- | --- | --- | --- | --- | --- | --- | --- | --- |
| 9 | Analysis of Nigerians with apparently sporadic Parkinson disease for mutations in LRRK2, PRKN and ATXN3 (2008). | Genetics | Case-control  PD=57 | Nigeria | UK  USA Portugal  Nigeria | Okubadejo et al | PLoS ONE  3.2 | Grant NS41723-01A1 from the National Institutes of Health. |
| 10 | A founding LRRK2 haplotype shared by Tunisian, US, European and Middle Eastern families with Parkinson's disease (2008). | Genetics | Prospective  Case-control  PD=132 | Tunisia | USA  UK  Tunisia | Warren et al | Parkinsonism  and related  disorders  4.8 | Not reported |
| 11 | Is the common LRRK2 G2019S mutation related to dyskinesias in North African Parkinson disease? (2008). | Genetics | Prospective  Case-control  PD=136 | Algeria Morocco Tunisia Libya | France | Lesage et al | Neurology  9.9 | Agence Nationale de la Recherche (ANR-05- NEUR-019). |
| 12 | Molecular analysis of the parkin gene in South African patients diagnosed with Parkinson's disease (2009). | Genetics | Prospective  Case-control  PD=91 | South Africa | None | Bardien et al | Parkinsonism  and related  disorders  4.8 | South African Medical Research Council, Harry and Doris Crossley Foundation and University of Stellenbosch, South Africa. |

Table 1(contd): Summary of included studies

| Study ID | Title and year | Theme | Design of study | African country(ies) defined by PD Cohort | Collaborating countries | First Author’s name | Journal and impact factor  2022 | Source of funding |
| --- | --- | --- | --- | --- | --- | --- | --- | --- |
| 13 | ATP13A2 variability in Parkinson disease (2009). | Genetics | Prospective  Case-control  PD=89 probands | Tunisia | UK  Canada | Vilarino-Guell et al | Human Mutation  4.7 | GlaxoSmithKline: (patient recruitment and clinical data collection). Molecular genetic analysis was supported by Neurogenetic Core of the Morris K. Udall Center, National Institute of Neurological Disorders and Stroke P50 NS40256 |
| 14 | A new complex homozygous large rearrangement of the PINK1 gene in a Sudanese family with early onset Parkinson's disease (2009). | Genetics | Prospective  Case report  PD=3 | Sudan | France Saudi Arabia | Cazeneuve et al | Neurogenetics  3.0 | Not reported |
| 15 | Identification of a novel functional deletion variant in the 5'-UTR of the DJ-1 gene (2009) | Genetics | Prospective  PD=88 | South Africa | None | Keyser et al | BMC Medical Genetics  2.0 | South African Medical Research Council, Harry and Doris Crossley Foundation and University of Stellenbosch, South Africa. |

Table 1(contd): Summary of included studies

| Study ID | Title and year | Theme | Design of study | African country(ies) defined by PD Cohort | Collaborating country | First Author’s name | Journal and impact factor  2022 | Source of funding |
| --- | --- | --- | --- | --- | --- | --- | --- | --- |
| 16 | A comparative study of LRRK2, PINK1 and genetically undefined familial Parkinson's disease (2010). | Genetics | Prospective  Cohort  PD=231 | Tunisia | USA  Poland  UK | Nishioka et al | Journal of Neurology, Neurosurgery and Psychiatry  13.6 | GlaxoSmithKline(patient recruitment and clinical data collection). Neurogenetic Core of a Morris K Udall Center, National Institute of Neurological Disorders and Stroke P50 NS40256 (Statistical analysis). Eli-Lilly scholarship and Herb Geist gift for Lewy body research (KN). Swiss National Science Foundation(CW) (PASMP3-123 268/1) |
| 17 | LRRK2 G2019S mutation: frequency and haplotype data in South African Parkinson's disease patients (2010). | Genetics | Case-control | South Africa (SA) | France | Bardien et al | Journal of Neural Transmission  3.5 | South Africa Medical Research Council, Harry and Doris Crossley Foundation,  University of Stellenbosch, SA and Agence Nationale de la Recherche,France |
| 18 | Assessing the prevalence of PINK1 genetic variants in South African patients diagnosed with early- and late-onset Parkinson's disease (2010). | Genetics | Prospective  Case-control  PD=154 | South Africa (SA) | France | Keyser et al | Biochemical and Biophysical Research Communications  3.7 | South Africa Medical Research Council, Harry and Doris Crossley Foundation, University of Stellenbosch, SA and Agence Nationale de la Recherche, France (ANR-05-NEUR-019 |

Table 1(contd): Summary of included studies

| Study ID | Title and year | Theme | Design of study | African country(ies) defined by PD Cohort | Collaborating country | First Author’s name | Journal and impact factor  2022 | Source of funding |
| --- | --- | --- | --- | --- | --- | --- | --- | --- |
| 19 | LRRK2 G2019S mutation in Parkinson's disease: a neuropsychological and neuropsychiatric study in a large Algerian cohort (2010). | Genetics | Case-control  PD=71 | Algeria | France | Belarbi et al | Parkinsonism  and related  disorders  4.8 | Ministère de la Santé, de la Population et de la Réforme Hospitalière and the Ministère de l’Enseignement Supérieur et de la Recherche Scientifique, Algeria |
| 20 | Genetic variation of the mitochondrial complex I subunit NDUFV2 and Parkinson's disease (2010). | Genetics | Case-control  PD=238 | Tunisia | USA | Nishioka et al | Parkinsonism  and related  disorders  4.8 | Morris K. Udall Center at Mayo Clinic Jacksonville, National Institute of Neurological Disorders and Stroke P50 NS40256 and R21 NS64885 and Michael J Fox Foundation for Parkinson’s Research. KN was supported by an Eli-Lilly scholarship and Herb Geist gift for Lewy body research |
| 21 | Glucocerebrosidase mutations are not a common risk factor for Parkinson disease in North Africa (2010). | Genetics | Prospective  Case-control  PD=273 | Tunisia | UK  USA | Nishioka et al | Neuroscience  Letters  2.1 | Same as No 20-GSK, Morris K. Udall Center, National Institute of Neurological Disorders and Stroke P50 NS40256 and MJFF. KN Eli-Lilly scholarship and Herb Geist gift for Lewy body research |

Table 1(contd): Summary of included studies

| Study ID | Title and year | Theme | Design of study | African country(ies) defined by PD Cohort | Collaborating countries | First Author’s name | Journal and impact factor  2022 | Source of funding |
| --- | --- | --- | --- | --- | --- | --- | --- | --- |
| 22 | Analysis of exon dosage using MLPA in South African Parkinson's disease patients (2010). | Genetics | Prospective  Cohort  PD=88 | South Africa | None | Keyser et al | Neurogenetics  3.0 | South African Medical Research Council, Harry and Doris Crossley Foundation, and University of Stellenbosch, South Africa. |
| 23 | Parkinson's disease-related LRRK2 G2019S mutation results from independent mutational events in humans (2010) | Genetics | Prospective  Case-control  PD=67(Africans) | Algeria Morocco Tunisia Libya | Turkey  Japan  South Africa | Lesage et al | Human Molecular Genetics  5.1 | Agence Nationale de la Recherche (ANR-05-NEUR-019). |

Table 1(contd): Summary of included studies

| Study ID | Title and year | Theme | Design of study | African country(ies) defined by PD Cohort | Collaborating countries | First Author’s name | Journal and impact factor  2022 | Source of funding |
| --- | --- | --- | --- | --- | --- | --- | --- | --- |
| 24 | Comprehensive sequencing of the LRRK2 gene in patients with familial Parkinson's disease from North Africa (2010). | Genetics | Prospective  Case-control  PD=259 | Tunisia | USA  Poland  UK  Switzerland | Jasinska-Myga et al | Movement Disorders  9.6 | Medical University of Silesia, Poland & Polish Foundation for Development of Neurology, Degenerative and Cerebrovascular Diseases & Pacific Alzheimer Research Foundation (PARF) (BJM & JF) grant C06-01. Swiss National Science Foundation (PASMP3-123268/1) CW. Morris K. Udall Center of Excellence for PD Research (P50-NS40256-MJF and Micheal J Fox Foundation (also OAR), GlaxoSmithKline, Institut National de Neurologie, La Rabta Tunis, Tunisia. LIP & RAG were supported by GlaxoSmithKline. RAG, MK, RA, SBY, SBS, MZ, GEE, FH supported by Ministry of Public Health in Tunisia. |

Table 1(contd): Summary of included studies

| Study ID | Title and year | Theme | Design of study | African country(ies) defined by PD Cohort | Collaborating country | First Author’s name | Journal and impact factor | Source of funding |
| --- | --- | --- | --- | --- | --- | --- | --- | --- |
| 25 | Low disease risk in relatives of north African LRRK2 Parkinson disease patients (2010). | Genetics | Cross-sectional  PD=236 | Algeria | France | Troiano et al | Neurology  9.9 | ANR (ANR-05- NEUR-O19) and INSERM (A05169DS). |
| 26 | Identification of Parkinson's disease candidate genes using CAESAR and screening of MAPT and SNCAIP in South African Parkinson's disease patients (2011) | Genetics | Case-control  PD=154 | South Africa (SA) | None | Keyser et al | Journal of Neural Transmission  3.5 | South Africa Medical Research Council, Harry and Doris Crossley Foundation,  University of Stellenbosch, SA and Agence Nationale de la Recherche,France |
| 27 | G2019S mutation of the leucine-rich repeat kinase 2 gene in a cohort of Egyptian patients with Parkinson's disease (2011). | Genetics | Cohort  PD=133  Controls=87 | Egypt | None | Hashad et al | Genetic testing and Molecular Biomarkers  1.7 | Not reported |
| 28 | Cognitive dysfunction in Tunisian LRRK2 associated Parkinson's disease (2011) | Genetics | Case-control  PD=110 | Tunisia | None | Ben Sassi et al | Parkinsonism  and related  disorders  4.8 | Not reported |
| 29 | Mutations in the parkin gene are a minor cause of Parkinson's disease in the South African population (2012) | Genetics | Cohort  PD=229 | South Africa | None | Haylett et al | Parkinsonism  and related  disorders  4.8 | South Africa (SA) Medical research council, Harry & Doris Crossley foundation & University of Stellenbosch, SA. |

Table 1(contd): Summary of included studies

| Study ID | Title and year | Theme | Design of study | African country(ies) defined by PD Cohort | Collaborating countries | First Author’s name | Journal and impact factor  2022 | Source of funding |
| --- | --- | --- | --- | --- | --- | --- | --- | --- |
| 30 | Analysis of LRRK2, SNCA, Parkin, PINK1, and DJ-1 in Zambian patients with Parkinson's disease (2012). | Genetics | Cohort  PD=39 | Zambia | Netherlands | Yonova-Doing et al | Parkinsonism  and related  disorders  4.8 | “International Parkinson Fonds”-The Netherlands, and Netherlands Organisation for Scientific research (NWO, VIDI grant) to VB. |
| 31 | Patient-control association study of the Leucine-Rich repeat kinase 2 (LRRK2) gene in South African Parkinson's disease patients (2013) | Genetics | Case-control  PD=205 | South Africa | Canada  USA | Bardien et al | Movement Disorders  9.6 | Michael J Fox foundation, Mayo clinic Morris K. Udall Parkinson’s disease research center of excellence & South African medical Research Council |
| 32 | STX6 rs1411478 is not associated with increased risk of Parkinson's disease (2013) | Genetics | Case-control  PD=146 | Tunisia | Canada  Norway  Taiwan | Trinh et al | Parkinsonism  and related  disorders  4.8 | Canada excellence research chairs program, province of British Columbia. (MJF), Leading Edge Endowment Funds provided by the Province of British Columbia, LifeLabs, and Genome BC support the Dr. Donald Rix BC Leadership Chair (MJF), and the Cundhill Foundation (MJF). |

Table 1(contd): Summary of included studies

| Study ID | Title and year | Theme | Design of study | African country(ies) defined by PD Cohort | Collaborating countries | First Author’s name | Journal and impact factor  2022 | Source of funding |
| --- | --- | --- | --- | --- | --- | --- | --- | --- |
| 33 | Screening of two indel polymorphisms in the 5'UTR of the DJ-1 gene in South African Parkinson's disease patients (2014). | Genetics | Prospective  Case-control  PD=402 | South Africa | None | Glanzmann et al | Journal of Neural Transmission  3.5 | South African Medical Research Council, Harry Crossley Foundation and Stellenbosch University. BG is supported by a National Research Foundation Innovation Doctoral Scholarship. |
| 34 | A comparative study of Parkinson's disease and leucine-rich repeat kinase 2 p.G2019S parkinsonism (2014) | Genetics | Cohort  PD=778 | Tunisia | Canada  USA  UK | Trinh et al | Neurobiology of Aging  4.6 | Michael J. Fox foundation and NIH. The Canadian excellence research chairs program, the Cundill foundation. |
| 35 | LRRK2 parkinsonism in Tunisia and Norway: A comparative analysis of disease penetrance (2014). | Genetics | Case-control  PD=582 | Tunisia | Canada  Norway  Taiwan | Hentati et al | Neurology  9.9 | Michael J.Fox Foundation, Canada excellence research chairs program, Canada Institute of Health Research and the Cundil Foundation, Province of British Columbia. |
| 36 | EIF4G1 R1205H and VPS35 D620N mutations are rare in Parkinson's disease from South Africa (2014) | Genetics | Case-control  PD=418 | South Africa | None | Blanckenberg et al | Neurobiology of Aging  4.6 | South African Medical Research Council, National Research Foundation and Harry Crossley Foundation and University of Stellenbosch. |

Table 1(contd): Summary of included studies

| Study ID | Title and year | Theme | Design of study | African country(ies) defined by PD Cohort | Collaborating countries | First Author’s name | Journal and impact factor | Source of funding |
| --- | --- | --- | --- | --- | --- | --- | --- | --- |
| 37 | Mitochondrial impairment observed in fibroblasts from South African Parkinson's disease patients with parkin mutations (2014). | Genetics | Prospective  Case-control  PD=3 | South Africa | None | van der Merwe et al | Biochemical and Biophysical Research communications  3.5 | Not reported |
| 38 | Genetic Analysis of Leucin-Rich Repeat Kinase 2 (LRRK2) G2019S Mutation in a Sample of Egyptian Patients with Parkinson's Disease, a Pilot Study (2014). | Genetics | Case-control  PD=69 | Egypt | Germany | El Desoky et al | British Journal of Medicine & Medical Research  2.0 | Not reported |
| 39 | Exonic rearrangements in the known Parkinson's disease-causing genes are a rare cause of the disease in South African patients (2016). | Genetics | Case-control  PD=210 | South Africa | None | van der Merwe et al | Neuroscience Letters  2.1 | South African Medical Research Council, the Harry Crossley Foundation, the National Research Foundation, and the University of Stellenbosch, South Africa. The SA MRC Centre for TB Research, DST/NRF Centre of Excellence for Biomedical TB Research. |

Table 1(contd): Summary of included studies

| Study ID | Title and year | Theme | Design of study | African country(ies) defined by PD Cohort | Collaborating countries | First Author’s name | Journal and impact factor  2022 | Source of funding |
| --- | --- | --- | --- | --- | --- | --- | --- | --- |
| 40 | A Novel Homozygous p.L539F Mutation Identified in PINK1 Gene in a Moroccan Patient with Parkinsonism (2016). | Genetics | Prospective/  Cohort  PD=1 | Morocco | None | Ben El Haj et al | BioMed Research International  3.4 | Centre National de Recherche Scientifique et Technique” (CNRST), “Minist`ere de l’Enseignement Sup´erieur, de la Recherche Scientifique et de la Formation des Cadres” (MESRSFC), and Mohammed V University in Rabat (UM5R), Morocco. |
| 41 | Greater improvement in LRRK2 G2019S patients undergoing Subthalamic Nucleus Deep Brain Stimulation compared to non-mutation carriers (2016). | Genetics | Case-control  PD=107 | Algeria | France | Sayad et al | BMC Neuroscience  3.2 | Not reported |
| 42 | A molecular analysis of the GBA gene in Caucasian South Africans with Parkinson's disease (2017) | Genetics | Case-control | South Africa | Netherlands | Barkhuizen et al | Molecular Genetics & Genomic Medicine  2.1 | National Research Foundation South Africa. |

Table 1(contd): Summary of included studies

| Study ID | Title and year | Theme | Design of study | African country(ies) defined by PD Cohort | Collaborating countries | First Author’s name | Journal and impact factor  2022 | Source of funding |
| --- | --- | --- | --- | --- | --- | --- | --- | --- |
| 43 | Using KASP technique to screen LRRK2 G2019S mutation in a large Tunisian cohort (2017). | Genetics | Case-control  PD=250 | Tunisia | None | Landoulsi et al | BMC Medical Genetics  2.0 | Tunisian Ministry of Higher Education and Scientific Research. Z.L has support within the framework of MOBIDOC Postdoc as specified in the PASRI program, funded by the European Union and managed by the ANPR. |
| 44 | Evidence for prehistoric origins of the G2019S mutation in the North African Berber population (2017). | Genetics | Case-control  PD=57 | Morocco | France | Ben El Haj et al | PLoS ONE  3.2 | “Centre  National de Recherche Scientifique et Technique”  (CNRST) of “Ministère de l'Enseignement  SupeÂrieur, de la Recherche Scientifique et de la  Formation des Cadres” (MESRSFC) and the  Mohammed V University in Rabat, (UM5R)  Morocco. |
| 45 | LRRK2 G2019S Mutation: Prevalence and Clinical Features in Moroccans with Parkinson's Disease (2017) | Genetics | Case-control  PD=100 | Morocco | None | Bouhouche et al | Parkinson's Disease  3.1 | Novartis pharma, centre national de recherche Scientifique et technique. |

Table 1(contd): Summary of included studies

| Study ID | Title and year | Theme | Design of study | African country(ies) defined by PD Cohort | Collaborating countries | First Author’s name | Journal and impact factor  2022 | Source of funding |
| --- | --- | --- | --- | --- | --- | --- | --- | --- |
| 46 | PTRHD1 loss-of-function mutation in an African family with parkinsonism and intellectual disability (2018). | Genetics | Observational cross-sectional | South Africa | Netherlands | Kuipers et al | Movement Disorders  9.6 | Stichting Parkinson Fonds (The Netherlands) to V.B., by the National Research Foundation of South Africa.  South African Medical Research Council |
| 47 | LRRK2 G2019S Parkinson's disease with more benign phenotype than idiopathic (2018). | Genetics | Case-control  PD=107 | Tunisia | France | Ben Romdhan et al | Acta Neurologica Scandinavica  3.9 | Not Reported |
| 48 | Leucine rich repeat kinase 2 (LRRK2) GLY2019SER mutation is absent in a second cohort of Nigerian Africans with Parkinson disease (2018) | Genetics | Case-control  PD=126 | Nigeria | UK | Okubadejo et al | PLoS ONE  3.2 | Intramural Research Program of the National Institute on Aging, National Institutes of Health, Dept. of Health and Human Services NUO received a University of Lagos Central Research Committee Grant to fund recruitment of study participants (clinical data collection). Funders website: www.unilag.edu.ng. Grant ID: CRC 11-004-5125 |

Table 1(contd): Summary of included studies

| Study ID | Title and year | Theme | Design of study | African country(ies) defined by PD Cohort | Collaborating countries | First Author’s name | Journal and impact factor 2022 | Source of funding |
| --- | --- | --- | --- | --- | --- | --- | --- | --- |
| 49 | Interleukin-18 promoter polymorphisms and idiopathic Parkinson disease: an Egyptian study (2019) | Genetics | Case-control  PD=30 | Egypt | None | Fahmy et al | Acta Neurologica Belgica  1.3 | Personally funded by authors |
| 50 | PINK1 Type of Early Onset Parkinson's Disease (EOPD) in Sudanese Patients, 2018 (2019). | Genetics | Prospective Cross-sectional  PD=31 | Sudan | None | Ibrahim et al | Sudan Journal of Medical Sciences  0.4 | Not reported |
| 51 | Frequency of the LRRK2 G2019S mutation in South African patients with Parkinson's disease (2019). | Genetics | Case-control  PD=647 | South Africa | None | du Toit et al | Neurogenetics  3.0 | National Research Foundation of South Africa & the South African Medical Research Council. NRF-DST Centre of Excellence for Biomedical Tuberculosis Research; Division of Molecular  Biology and Human Genetics, Faculty of Medicine and Health  Sciences, Stellenbosch University, Cape Town |

Table 1(contd): Summary of included studies

| Study ID | Title and year | Theme | Design of study | African country(ies) defined by PD Cohort | Collaborating countries | First Author’s name | Journal and impact factor | Source of funding |
| --- | --- | --- | --- | --- | --- | --- | --- | --- |
| 52 | Screening of the glucocerebrosidase (GBA) gene in South Africans of African ancestry with Parkinson's disease (2019). | Genetics | Case-control  PD=30 | South Africa (SA) | USA | Mahungu et al | Neurobiology of Aging  4.6 | National Research Foundation of SA, the SA Medical Research Council & Stellenbosch University. NRF-DST Centre of Excellence for Biomedical Tuberculosis Research; Division of Molecular Biology and Human Genetics, Faculty of Medicine & Health Sciences, Stellenbosch University, NIH, Michael J. Fox Foundation, the American Parkinson Disease Association (APDA) Mayo Clinic Information and Referral Center |

Table 1(contd): Summary of included studies

| Study ID | Title and year | Theme | Design of study | African country(ies) defined by PD Cohort | Collaborating countries | First Author’s name | Journal and impact factor  2022 | Source of funding |
| --- | --- | --- | --- | --- | --- | --- | --- | --- |
| 53 | Targeted next-generation sequencing identifies novel variants in candidate genes for Parkinson's disease in Black South African and Nigerian patients (2020). | Genetics | Cross-sectional  PD=47 | South Africa (SA)  Nigeria | USA | Oluwole et al | BMC Medical Genetics  2.0 | National Institute of Neurological Disorders and Stroke, and Fogarty International Center, the National Institutes of Health, USA, SA National Research Foundation, SA Medical Research Council. Faculty of Medicine and Health Sciences, Stellenbosch University,SA. SA Tuberculosis Bioinformatics Initiative (SATBBI). NIH grant for the Mayo Clinic Morris K. Udall Center of Excellence in Parkinson’s Disease Research. |
| 54 | PRKN-related familial Parkinson's disease: First molecular confirmation from East Africa (2020). | Genetics | Case series of a family  PD=3 | Tanzania | Netherlands | Dekker et al | Parkinsonism  and related  disorders  4.8 | No funding |
| 55 | Effects of glutathione S-transferase M1 and T1 deletions on PD risk among a North African population (2020) | Genetics | Prospective case-control  PD=64 | Tunisia | None | Rebai et al | Revue Neurologique  2.6 | Not reported |

Table 1(contd): Summary of included studies

| Study ID | Title and year | Theme | Design of study | African country(ies) defined by PD Cohort | Collaborating countries | First Author’s name | Journal and impact factor  2022 | Source of funding |
| --- | --- | --- | --- | --- | --- | --- | --- | --- |
| 56 | Association Between a Variable Number Tandem Repeat Polymorphism Within the DAT1 Gene and the Mesolimbic Pathway in Parkinson's Disease (2020). | Genetics | Cohort  PD=45 | South Africa (SA) | None | du Plessis et al | Frontiers of Neurology  4.0 | National Research Foundation of SA (Grant Number: 106052), SA Medical Research Council, the NRF-DST Center of Excellence for Biomedical Tuberculosis Research; SA Medical Research Council Center for Tuberculosis Research |
| 57 | Age at Onset of LRRK2 p.Gly2019Ser Is Related to Environmental and Lifestyle Factors (2020). | Genetics | Case Report  PD=199 | Tunisia | Luxemburg  Germany USA | Lüth et al | Movement Disorders  9.6 | DFG RU FOR2488 Protect Move (to C.K. and J.T.), MDS (to C.K.), Ger. Res. Foundation (to C.K.), BMBF (to C.K.), European Community to (C.K.), intramural funds from the Uni. of Luebeck (to C.K.), Peter and TE fellowship (to J.T.), CIHR Fellowship (to J.T.), Joachim Herz Stiftung Add-on fellowship (to J.T.) and Else Kroner Fresenius Stiftung (to J.T.). |

Table 1(contd): Summary of included studies

| Study ID | Title and year | Theme | Design of study | African country(ies) defined by PD Cohort | Collaborating countries | First Author’s name | Journal and impact factor  2022 | Source of funding |
| --- | --- | --- | --- | --- | --- | --- | --- | --- |
| 58 | Gene Panel Sequencing Identifies Novel Pathogenic Mutations in Moroccan Patients with Familial Parkinson Disease (2021). | Genetics | Cohort  PD=18 | Morocco | None | Smaili et al. | Journal of Molecular Neuroscience  3.4 | “Ministère de l’Enseignement Supérieur, de la Recherche Scientifique et de la Formation des Cadres” (MESRSFC) of Morocco and the “Centre National de Recherche Scientifique et Technique” (CNRST). |
| 59 | Negative screening for 12 rare LRRK2 pathogenic variants in a cohort of Nigerians with Parkinson's disease (2021) | Genetics | Case-control  PD=92 | Nigeria | UK | Rizig et al | Neurobiology of Aging  4.6 | University of Lagos Central Research Committee (grant -CRC 11-004–5125) for recruitment of study participants (NUO) and the University College London Global Challenge Research Grant (Award ID:177813) for DNA extraction and genotyping (MR). |

Table 1(contd): Summary of included studies

| Study ID | Title and year | Theme | Design of study | African country(ies) defined by PD Cohort | Collaborating countries | First Author’s name | Journal and impact factor | Source of funding |
| --- | --- | --- | --- | --- | --- | --- | --- | --- |
| 60 | Prioritization of candidate genes for a South African family with Parkinson’s disease using in-silico tools (2021). | Genetics | Cohort  PD=11 | South Africa (SA) | None | Sebate et al. | PLoS ONE  3.2 | SB and JC received support from the National Research Foundation of SA (Grant Number: 106052) and SA Medical Research Council (Self-Initiated Research Grant). RC and AC were supported by the SA Research Chairs Initiative of the Dept of Science and Technology and National Research Foundation (NRF) of SA (award number UID 64751). |
| 61 | Early-Onset Parkinson Disease Screening in Patients from Nigeria (2021) | Genetics | Cohort  PD=15 | Nigeria | USA | Milanowski et al | Frontiers of Neurology  4.0 | American Parkinson Disease Association (APDA) Mayo Clinic Information and Referral Center/Polish National Agency for Academic Exchange Iwanowska's Fellowship/National Institutes of Health /National Institute of Neurological Disorders and Stroke (NINDS) |

Table 1(contd): Summary of included studies

| Study ID | Title and year | Theme | Design of study | African country(ies) defined by PD Cohort | Collaborating countries | First Author’s name | Journal and impact factor | Source of funding |
| --- | --- | --- | --- | --- | --- | --- | --- | --- |
| 62 | Study of cognitive impairment and genetic polymorphism of SLC41A1 (rs11240569 allele) in Parkinson’s disease in Upper Egypt: case-control study (2021). | Genetics | Case-control  PD=41 | Egypt | None | El-tallawy et al. | The Egyptian Journal of Neurology, Psychiatry and Neurosurgery  0.7 | Not reported |
| 63 | APOE E4 is associated with impaired self-declared cognition but not disease risk or age of onset in Nigerians with Parkinson’s disease (2022). | Genetics | Case-control | Egypt | UK  USA | Okubadejo et al | Nature Parkinson’s Journal  9.3 | R.A. is supported by: US NIH/ NHGRI & UK Royal Society /African Academy of Sciences M.R.: UCL Grand Challenges Small Grants & Michael J Fox Foundation (MJFF) Genetic Diversity (GD) in PD 2019. H.H.: MJFF GD in PD. N.U.O: MJFF GD in PD 2019 & TETFund National Research Fund (NRF) 2019. S.B.-C., C.B. and A.S. are supported by the IRP, National Institute on Aging, NIH and US Department of Health and Human Services. |

Table 1(contd): Summary of included studies

| Study ID | Title and year | Theme | Design of study | African country(ies) defined by PD Cohort | Collaborating countries | First Author’s name | Journal and impact factor  2022 | Source of funding |
| --- | --- | --- | --- | --- | --- | --- | --- | --- |
| 64 | Heart rate variability and sympathetic skin  response for the assessment of autonomic  dysfunction in leucine-rich repeat kinase 2  associated Parkinson’s disease (2022). | Genetics | Retrospective  Cross-sectional | Tunisia | None | Nasri et al | Neurophysiologie Clinique  3.7 | Not reported |
| 65 | Methylation of alpha-synuclein in a Sudanese cohort (2022) | Genetics | Cohort  Case-control | Sudan | France  Germany | Bakhit et al | Parkinsonism  and related  disorders  4.8 | None |
| 66 | Increased blood-derived mitochondrial DNA copy number in African ancestry individuals with Parkinson’s disease (2022) | Genetics | Case-control | South Africa | None | Muller-Nedebock et al | Parkinsonism  and related  disorders  4.8 | Supported in part by the South African Medical Research Council [Self-Initiated Research Grant]; the National Research Foundation of South Africa [grant numbers: 106052; 120719; 123256; and 129249]; and the Harry Crossley Foundation. |

Table 1(contd): Summary of included studies

| Study ID | Title and year | Theme | Design of study | African country(ies) defined by PD Cohort | Collaborating countries | First Author’s name | Journal and impact factor  2022 | Source of funding |
| --- | --- | --- | --- | --- | --- | --- | --- | --- |
| 67 | Parkinson's disease: The prevalence of the LRRK2-G2019S mutation among the population of the South-West of Morocco (2022). | Genetics | Case-control | Morocco | None | Wahmane et al | Gene Reports  1.51 | This research was supported by the Laboratory of Cell Biology and Molecular Genetics, Faculty of Sciences, Ibn Zohr University. |
| 68 | Frequency and predictors of autonomic dysfunction in PD: A study of African Patients in Lagos, Nigeria (2004). | Clinical features | Prospective  PD=33  Control=33 | Nigeria | None | Okubadejo et al | The Nigerian Postgraduate Medical Journal  0.6 | None |
| 69 | Clinical Features of Parkinson Disease Patients with Homozygous Leucine-Rich Repeat Kinase 2 G2019S Mutations (2006) | Clinical features | Case-control  PD=209 | Tunisia | France | Ishihara L et al | Archives of Neurology  (now JAMA)  29.9 | GlaxoSmithKline |
| 70 | Cognitive Dysfunction in Nigerians with Parkinson’s Disease (2008) | Clinical features | Case-control  PD=52  Controls=50 | Nigeria | None | Akinyemi et al | Movement Disorders  9.6 | None |
| 71 | Motor and non-motor features of Parkinson’s disease (2008) | Clinical features | Prospective  PD=42 | Tanzania | None | Matuja et al | East African Medical Journal  0.03 | None |

Table 1(contd): Summary of included studies

| Study ID | Title and year | Theme | Design of study | African country(ies) defined by PD Cohort | Collaborating countries | First Author’s name | Journal and impact factor  2022 | Source of funding |
| --- | --- | --- | --- | --- | --- | --- | --- | --- |
| 72 | Non-motor symptoms in a prevalent population with Parkinson’s disease in Tanzania (2009). | Clinical features | Prospective  PD=33 | Tanzania | UK | Dotchin CL et al | Parkinsonism  and related  disorders  4.8 | UK Parkinson’s Disease Society and Northumbria Healthcare NHS Trust. |
| 73 | Disorders of basal ganglia at the university teaching hospital Lusaka (2009). | Clinical features | Retrospective  PD=50 | Zambia | None | Atadzhanov et al | Medical Journal of Zambia  NA | None |
| 74 | Clinical presentation of Parkinson's disease among Sudanese patients (2009) | Clinical features | Descriptive/ prospective/ Cross- sectional  PD=94 | Sudan | None | Khalid K et al | Sudan Journal of Medical Sciences  0.4 | None |
| 75 | Clinical profile of parkinsonism and Parkinson's disease in Lagos, Southwestern Nigeria (2010). | Clinical features | Prospective  PD=98 | Nigeria | None | Okubadejo et al | BMC Neurology  2.2 | None |
| 76 | Plasma Homocysteine level and its Relationship to Clinical Profile in Parkinson’s Disease Patients at the Lagos University Teaching Hospital (2011). | Clinical features | Prospective  Case-control  PD=40  Control=40 | Nigeria | None | Ojo et al | West African Journal of Medicine  0.2 | None |

Table 1(contd): Summary of included studies

| Study ID | Title and year | Theme | Design of study | African country(ies) defined by PD Cohort | Collaborating countries | First Author’s name | Journal and impact factor | Source of funding |
| --- | --- | --- | --- | --- | --- | --- | --- | --- |
| 77 | Pneumophonic Coordination Impairments in Parkinsonian Dysarthria: Importance of Aerodynamic Parameters Measurements (2012). | Clinical features | Prospective  Case-control  PD=24  Controls=50 | Senegal | France | Moustapha SM et al | West African Journal of Medicine  0.2 | Grant from the SAC of the Embassy of France in Dakar and the support of France Parkinson Association. |
| 78 | Assessment of speech in neurological disorders: Development of a Swahili screening test (2012). | Clinical features | Prospective  Case-control  PD=26 | Tanzania | UK | Miller et al | South African Journal of Communication Disorders  0.9 | British Academy UK-Africa Academic Partnerships. |
| 79 | Frequency of cognitive impairment and depression in Parkinson's disease: A preliminary case-control study (2012). | Clinical features | Case-control  PD=40  Controls=40 | Nigeria | None | Ojo et al | Nigerian Medical Journal  0.7 | Not reported |
| 80 | Gastrointestinal complications in newly diagnosed Parkinson’s disease: A case-control study | Clinical features | Prospective  PD=80 | Nigeria | None | Owolabi LF et al | Journal of Neurosciences in Rural Practice  0.7 | Not reported |
| 81 | Depression among Patients with Parkinson's Disease in a Nigerian Tertiary Hospital (2013). | Clinical features | Prospective  Case-control  PD=36 | Nigeria | None | Okunoye et al | The Nigerian Health Journal  NA | None |

Table 1(contd): Summary of included studies

| Study ID | Title and year | Theme | Design of study | African country(ies) defined by PD Cohort | Collaborating countries | First Author’s name | Journal and impact factor | Source of funding |
| --- | --- | --- | --- | --- | --- | --- | --- | --- |
| 82 | Autonomic function in a prevalent Tanzanian population with Parkinson’s disease and its relationship to disease duration and 5-year mortality (2013). | Clinical features | Prospective  Cohort  PD=29 | Tanzania | UK | Aris et al | BMC Research Note  2.15 | Not reported |
| 83 | Somatosensory and Motor Systems Affect Postural Stability in Parkinson’s Disease Patients (2013). | Clinical features | Prospective Case-control  PD=20  Controls=20 | Egypt | None | Darwish et al | The Egyptian Journal of Neurology, Psychiatry and Neurosurgery  0.7 | Not reported |
| 84 | Neuropsychiatric symptoms in Nigerian patients with Parkinson's disease (2013). | Clinical features | Prospective  Case-control  PD=50  Controls=50 | Nigeria | None | Ojagbemi et al | Acta Neurologica Scandinavica  3.9 | Not reported |
| 85 | Relationship Between Cognitive Dysfunction and Behavioural Symptoms in Nigerian Patients with Parkinson’s Disease No Dementia (2013). | Clinical features | Prospective  Case-control  PD=50 | Nigeria | None | Ojagbemi et al | Journal of Parkinson’s Disease  5.5 | Not reported |
| 86 | Comparison of the clinical profile of Parkinson's disease between Spanish and Cameroonian Cohorts (2014). | Clinical features | Cohort  PD=37 | Cameroun | Spain | Cubo et al | Journal of the Neurological Sciences  3.1 | Research Intramural Program of the Carlos III Institute of Health (EPY1271/05). |

Table 1(contd): Summary of included studies

| Study ID | Title and year | Theme | Design of study | African country(ies) defined by PD Cohort | Collaborating countries | First Author’s name | Journal and impact factor | Source of funding |
| --- | --- | --- | --- | --- | --- | --- | --- | --- |
| 87 | Phenotypic Characteristics of Zambian patients with Parkinson's Disease (2014). | Clinical features | Prospective  Case-control  PD=46  Controls=46 | Zambia | None | Atadzhonov et al | Medical Journal of Zambia  0.13 | Not reported |
| 88 | Preliminary Investigation of Risk Factors Causing Dyskinesias in Parkinson’s Disease in South Africa (2014). | Clinical features | Prospective/  Retrospective  PD=43 | South Africa | None | Gaida et al | Tropical Journal of Pharmaceutical Research | The Nelson Mandela Metropolitan University (NMMU) and the National Research Foundation (NRF) |
| 89 | Gastrointestinal complications in newly diagnosed Parkinson’s disease: A case-control study (2014). | Clinical features | Prospective  Case-control  PD=80  Control=80 | Nigeria | None | Owolabi et al | Tropical Gastroenterology | Not reported |
| 90 | Profile of idiopathic Parkinson’s disease in Moroccan patients (2014) | Clinical features | Retrospective  PD=117 | Morocco | None | Regragul et al | International Archives of Medicine | Not reported |
| 91 | The modern pre-levodopa era of Parkinson’s Disease: insights into motor complications from sub-Saharan Africa (2014). | Clinical features | Prospective  PD=91 | Ghana | Italy | Cilia et al | Brain: A Journal Neurology | Fondazione Grigioni per il Morbo di Parkinson and Regione Lombardia |

Table 1(contd): Summary of included studies

| Study ID | Title and year | Theme | Design of study | African country(ies) defined by PD Cohort | Collaborating countries | First Author’s name | Journal and impact factor | Source of funding |
| --- | --- | --- | --- | --- | --- | --- | --- | --- |
| 92 | Non motor features of Parkinson disease patients attending neurology clinic at tertiary institution in Nigeria (2014). | Clinical features | Prospective  Case-control  PD=36 | Nigeria | None | Okunoye et al | The Nigerian Health Journal  NA | None |
| 93 | The Clinical Profile Of Idiopathic Parkinson’s Disease In A South African Hospital Complex - The Influence Of Ethnicity And Gender (2015) | Clinical features | Prospective  PD=50 | South Africa | None | Smith et al | African Journal of Neurological Sciences  3.1 | Not reported |
| 94 | Sleep quality assessment in 35 Parkinson’s disease patients in the Fann Teaching Hospital, Dakar,Senegal(2015). | Clinical features | Prospective  PD=35 | Senegal | None | Maiga et al | Revue neurologique  2.6 | Not reported |
| 95 | Levels of functional disability in elderly people in Tanzania with dementia, stroke and Parkinson’s disease (2015) | Clinical features | Prospective  PD=12 | Tanzania | UK | Kisoli A et al | Acta Neuropsychiatrica  4.5 | Dunhill Foundation, RCP, Peel Medical Research Trust, British Geriatric Society SpR start up grant, Academy of Medical Sciences (UK) Clinical Lecturer start up grant and Northumbria Healthcare NHS Foundation Trust. |

Table 1(contd): Summary of included studies

| Study ID | Title and year | Theme | Design of study | African country(ies) defined by PD Cohort | Collaborating countries | First Author’s name | Journal and impact factor | Source of funding |
| --- | --- | --- | --- | --- | --- | --- | --- | --- |
| 96 | Pulmonary function tests in patients with Parkinson’s disease: A case‑control study (2016). | Clinical features | Prospective  Case-control  PD=78  Controls=78 | Nigeria | None | Owolabi et al | Nigerian Journal of Clinical Practice  0.9 | Not reported |
| 97 | Falls and Their Associated Risks in Parkinson’s Disease Patients in Nigeria (2016) | Clinical features | Prospective  Case-control  PD=81 | Nigeria | None | Farombi et al | The Journal of Movement Disorders  4.2 | Not reported |
| 98 | Non motor symptoms as predictors of quality of life in Egyptian patients with PD: a cross sectional study using a culturally adapted 39-item PD questionnaire (2018). | Clinical features | Prospective  PD=97 | Egypt | None | Shalash et al | Frontier in Neurology  4.0 | Not reported |
| 99 | Clinical profile of Parkinson’s disease: Experience of Niger. (2018) | Clinical features | Retrospective  PD=25 | Niger | UK | Assadeck et al | Journal of Neurosciences in Rural Practice  0.7 | Not reported |
| 100 | Frequency and Clinical profile of Parkinson's disease and other Parkinsonian syndromes seen in the Department of Neurology at the Befelatanana Hospital Antananarivo (2019). | Clinical features | Retrospective  PD=67 | Madagascar | None | Rasaholiarison et al | Pan African Medical Journal  0.8 | Not reported |

Table 1(contd): Summary of included studies

| Study ID | Title and year | Theme | Design of study | African country(ies) defined by PD Cohort | Collaborating countries | First Author’s name | Journal and impact factor | Source of funding |
| --- | --- | --- | --- | --- | --- | --- | --- | --- |
| 101 | Profile of Non-motor Symptoms and the Association with the Quality of Life of Parkinson's Disease Patients in Nigeria (2019). | Clinical features | Prospective  Case-control  PD=105  Controls=105 | Nigeria | None | Arambabi et al | Nigerian Medical Journal  0.7 | Not reported |
| 102 | Clinical Profile of Parkinson`s Disease in Calabar, Southern Nigeria (2019) | Clinical features | Prospective  PD=42 | Nigeria | None | Oparah et al | Journal of Research in Basic & Clinical Sciences  NA | Not reported |
| 103 | Prevalence of pain in patients with Parkinson's disease in Addis Ababa, Ethiopia (2019). | Clinical features | Prospective  PD=103 | Ethiopia | None | Hirsi et al | Parkinsonism  and related  disorders  4.8 | Not reported |
| 104 | Impulse control disorders in Parkinson disease: A cross-sectional study in Morocco (2019) | Clinical features | Prospective  PD-125 | Morocco | None | El Otmani H et al | Revue neurologique  2.6 | Not reported |
| 105 | Prevalence of sleep disorders in Parkinson’s disease patients in two neurology referral hospitals in Ethiopia (2019) | Clinical features | Prospective  PD=155 | Ethiopia | USA | Melka D et al | BMC Neurology  2.2 | Not reported |

Table 1(contd): Summary of included studies

| Study ID | Title and year | Theme | Design of study | African country(ies) defined by PD Cohort | Collaborating countries | First Author’s name | Journal and impact factor | Source of funding |
| --- | --- | --- | --- | --- | --- | --- | --- | --- |
| 106 | Clinical series of Parkinson's disease in KwaZulu-Natal, South Africa: Retrospective chart review (2019). | Clinical features | Retrospective  PD=414 | South Africa | None | Amod FH et al | Journal of the Neurological Sciences  3.1 | Not reported |
| 107 | Prevalence and determinants of Fatigue among Parkinson’s disease patients in Ethiopia (2020) | Clinical features | Cross-sectional  PD=155 | Ethiopia | None | Melka D et al | Ethiopian Medical Journal  0.3 | Not reported |
| 108 | The Nigeria Parkinson Disease Registry: Process, Profile, and Prospects of a Collaborative Project. (2020). | Clinical features | Observational  N=578 | Nigeria | None | Ojo et al | Movement Disorders  9.6 | Not reported |
| 109 | Translation, Validation, Diagnostic Accuracy, and Reliability of Screening Questionnaire for Parkinsonism in Three African Countries (2020) | Clinical features | Cross-sectional  PD=159 | Nigeria  Egypt  Cameroon | USA  Spain | Shalash et al. | Journal of Parkinson's Disease  5.5 | Not reported |

Table 1(contd): Summary of included studies

| Study ID | Title and year | Theme | Design of study | African country(ies) defined by PD Cohort | Collaborating countries | First Author’s name | Journal and impact factor | Source of funding |
| --- | --- | --- | --- | --- | --- | --- | --- | --- |
| 110 | Validation of Parkinson’s Disease-Related Questionnaires in South Africa (2020) | Clinical features | Survey | South Africa | USA  UK | Nelson et al. | Hindawi  2.8 | University of the Witwatersrand School of Public Health (seed funding), and University of the Witwatersrand Faculty Research Grant. |
| 111 | Gender and Age Difference in Clinical Features and severity of Parkinson’s Disease: A Cross-Sectional Study in Southern Morocco (2020). | Clinical features | Cross-sectional  PD=180 | Morocco | None | Achbani et al. | Archives of Neuroscience  0.5 | This research was supported by the Laboratory of Cell Biology and Molecular Genetics, Faculty of Sciences of Agadir. |
| 112 | Natural history of motor symptoms in Parkinson’s disease and the long-duration response to levodopa (2020) | Clinical features | Cohort  PD=30 | Ghana  Zambia | Italy | Cilia et al | Brain  8.7 | This work was supported by the ‘Fondazione Grigioni per ilMorbo di Parkinson’, Milan, Italy, which was the main sponsor of the project since December 2008. |
| 113 | Non-motor symptoms in essential tremor, akinetic rigid and tremor-dominant subtypes of Parkinson’s disease. | Clinical features | Restrospective  PD=72  Controls=28 | Egypt | None | Shalash et al | PLoS ONE  3.2 | None |

Table 1(contd): Summary of included studies

| Study ID | Title and year | Theme | Design of study | African country(ies) defined by PD Cohort | Collaborating countries | First Author’s name | Journal and impact factor at  2022 | Source of funding |
| --- | --- | --- | --- | --- | --- | --- | --- | --- |
| 114 | A Cross-Sectional Comprehensive Assessment of the Profile and Burden of Non-motor Symptoms in Relation to Motor Phenotype in the Nigeria Parkinson Disease Registry Cohort (2021) | Clinical features | Cross-sectional  PD=825 | Nigeria | UK | Ojo et al. | Movement disorders clinical practice | None |
| 115 | A Rapid Motor Task-Based Screening Tool for Parkinsonism in Community-Based Studies (2021). | Clinical features | Cross-sectional  PD=57 | South Africa | USA  UK | Dlamini et al. | Movement Disorders  9.9 | National Institutes of Health–National Institute of Environmental Health Sciences (Grant Nos. R01ES025991, R01ES026891-S1, and K01ES028295). The funder had no role in the design and conduct of the study; collection, statistical analysis, or interpretation of the data; preparation, review, or approval of the manuscript; and the decision to publish these results. |

Table 1(contd): Summary of included studies

| Study ID | Title and year | Theme | Design of study | African country(ies) defined by PD Cohort | Collaborating countries | First Author’s name | Journal and impact factor | Source of funding |
| --- | --- | --- | --- | --- | --- | --- | --- | --- |
| 116 | No impact of confinement during COVID-19 pandemic on anxiety and depression in Parkinsonian patients (2021) | Clinical features | Prospective Cohort  PD=50 | Morocco | None | El Otmani et al. | Revue neurologique  2.6 | Not reported |
| 117 | Non-Motor Symptoms and Associated Factors in Parkinson's Disease Patients in Addis Ababa, Ethiopia: A Multicenter Cross-Sectional Study (2021). | Clinical features | Cross-sectional  Observational  PD=123 | Ethiopia | None | Ayele et al. | Ethiopian journal of health sciences  1.18 | Not reported |
| 118 | “Old people problems”, uncertainty and legitimacy: Challenges with diagnosing Parkinson's disease in Kenya (2021) | Clinical features | Interviews/  Surveys  PD=55 | Kenya | UK | Fothergill-Misbah et al. | Social science & medicine  4.6 | UKRI, Economic and Social Research Council (ESRC) Northern Ireland and North East Doctoral Training Partnership, United Kingdom (grant number [ES/J500082/1](https://www-sciencedirect-com.libproxy.ucl.ac.uk/science/article/pii/S0277953621004809#gs5)). |

Table 1(contd): Summary of included studies

| Study ID | Title and year | Theme | Design of study | African country(ies) defined by PD Cohort | Collaborating countries | First Author’s name | Journal and impact factor | Source of funding |
| --- | --- | --- | --- | --- | --- | --- | --- | --- |
| 119 | Prevalence of abnormal pulmonary functions in Parkinson’s disease and its correlation to the disease severity and quality of life (2021). | Clinical features | Case-control  PD=52 | Egypt | None | Elsherif et al. | Medical Sciences  2.3 | Not reported |
| 120 | Study of Unilateral Spatial Neglect in Parkinson’s patients (2021) | Clinical features | Prospective  PD=60  Controls=60 | Morocco | None | Ahmadou et al | Acta Neuropsychologica  0.7 | Not reported |
| 121 | Atypical parkinsonian syndromes in a North African tertiary referral center (2021). | Clinical features | Prospective  PD=464 | Tunisia | None | Nasri et al. | Brain and Behavior  2.7 | Not reported |
| 122 | Does antiretroviral therapy alter the course of Parkinson's disease in people living with HIV? (2021) | Clinical features | Prospective  PD=20 | South Africa | None | Amod et al. | Journal of Neurovirology  2.6 | Not reported |
| 123 | The potential role of 2D-speckle tracking echocardiography for detecting  left ventricular systolic dysfunction in patients with Parkinson’s disease: a case control study (2021) | Clinical features | Case-control  PD=40  Control=40 | Egypt | None | El Mokadem et al | Acta Cardiologica  1.8 | None |

Table 1(contd): Summary of included studies

| Study ID | Title and year | Theme | Design of study | African country(ies) defined by PD Cohort | Collaborating countries | First Author’s name | Journal and impact factor  2022 | Source of funding |
| --- | --- | --- | --- | --- | --- | --- | --- | --- |
| 124 | Assessment of postural instability in  Parkinson’s disease patients (2021). | Clinical features | Case-control  PD=15 | Egypt | None | Talaat et al. | Egyptian Journal of Otolaryngology | None |
| 125 | Screening for non‑motor symptoms  in Egyptian patients with Parkinson’s disease (2022) | Clinical features | Case-control  PD=50 | Egypt | None | Shaheen et al | The Egyptian Journal of Neurology, Psychiatry and Neurosurgery  0.7 | None |
| 126 | The effect of hypocalcemia on motor  symptoms of Parkinson’s disease (2022). | Clinical features | Case-control  PD=28 | Egypt | None | Emad et al. | The Egyptian Journal of Neurology, Psychiatry and Neurosurgery  0.7 | None |
| 127 | Optical coherence tomography in patients  with Parkinson’s disease (2022) | Clinical features | Case-control  PD=32  Controls=32 | Egypt | None | El-Kattan et al. | The Egyptian Journal of Neurology, Psychiatry and Neurosurgery  0.7 | None |
| 128 | Frequency and Factors Associated with Orthostatic Hypotension in  Individuals with Parkinson’s Disease: A Case-Control Observational  Study (2022) | Clinical features | Case-control  Observational  PD=53  Controls=53 | Ethiopia | None | Mengesha et al | Ethiopian journal of health sciences  1.18 | Not reported |

Table 1(contd): Summary of included studies

| Study ID | Title and year | Theme | Design of study | African country(ies) defined by PD Cohort | Collaborating countries | First Author’s name | Journal and impact factor  2022 | Source of funding |
| --- | --- | --- | --- | --- | --- | --- | --- | --- |
| 129 | Factors Associated with EEG Slowing in Individuals with Parkinson's  Disease (2022). | Clinical features | Cross-sectional  Observational  PD=40 | Ethiopia | None | Ayele et al. | Ethiopian journal of health sciences  1.18 | Yehuleshet Specialty Clinic |
| 130 | Baseline predictors of progression  of Parkinson’s disease in a sample of Egyptian  patients: clinical and biochemical (2022) | Clinical features | Prospective cohort study  PD=45 | Egypt | None | Helmy et al | The Egyptian Journal of Neurology, Psychiatry and Neurosurgery  0.7 | None |
| 131 | Sexual Dysfunction Among Egyptian  Idiopathic Parkinson’s Disease Patients (2022). | Clinical features | Case-control  PD=67  Controls=30 | Egypt | None | Elshamy et al. | Journal of Geriatric Psychiatry and Neurology | None |
| 132 | A 6-month longitudinal study on worsening of Parkinson’s disease during the COVID-19 pandemic (2022). | Clinical features | Retrospective  PD=50 | Egypt | None | Shalash et al | NPJ Parkinson’s disease  9.0 | Open access funding provided by The Science, Technology & Innovation Funding Authority (STDF) in cooperation with The Egyptian Knowledge Bank (EKB). |
| 133 | Epidemiology of Parkinson's disease in Benghazi, North-East Libya (1986). | Epidemiology | Cross-sectional  Hospital- based | Libya | India | Ashok PP et al | Clinical neurology and neurosurgery | Not reported |

Table 1(contd): Summary of included studies

| Study ID | Title and year | Theme | Design of study | African country(ies) defined by PD Cohort | Collaborating countries | First Author’s name | Journal and impact factor  2022 | Source of funding |
| --- | --- | --- | --- | --- | --- | --- | --- | --- |
| 134 | Comparison of the prevalence of Parkinson’s disease in black populations in the rural United States and in rural Nigeria: Door-to-door community studies (1988). | Epidemiology | Cross-sectional  Population-based | Nigeria | USA | Schoenberg BS et al | Neurology  9.9 | Not reported |
| 135 | Community-based study of neurological disorders in rural Central Ethiopia (1990) | Epidemiology | Cross-sectional  Population-based | Ethiopia | Sweden | Takle-Haimanot et al | Neuroepidemiology  3.2 | Sweddish Agency for Research Cooperation with Developing Countries and Addis Ababa University |
| 136 | Prevalence study of neurologic disorders in Kelibia (Tunisia) (1993). | Epidemiology | Cross-sectional  Population-based | Egypt | None | Romdhane NA et al | Neuroepidemiology  3.2 | Ministère de l'éducation et de la recherche scientifique/Ministère de la santé/Caisse Nationale De Retraite Et De Prevoyance Sociale/ FIC-NINCDS-WHO fellowship grant |
| 137 | Epidemiological Features of Degenerative Brain Diseases as They Occurred in Yaoundé Referral Hospitals over a 9-Year Period (2006). | Epidemiology | Retrospective Hospital-based | Cameroun | Australia  France | Kengne AP et al | Neuroepidemiology  3.2 | Not Reported |

Table 1(contd): Summary of included studies

| Study ID | Title and year | Theme | Design of study | African country(ies) defined by PD Cohort | Collaborating countries | First Author’s name | Journal and impact factor  2022 | Source of funding |
| --- | --- | --- | --- | --- | --- | --- | --- | --- |
| 138 | The Prevalence of Parkinson’s Disease in Rural Tanzania (2008). | Epidemiology | Cross-sectional  Hospital & Population-based | Tanzania | UK | Dotchin C et al | Movement Disorders  9.9 | UK Parkinson’s Disease Society and Northumbria Healthcare NHS Trust |
| 139 | Profile of Neurological admissions at the University of Nigeria Teaching Hospital Enugu (2010) | Epidemiology | Retrospective Hospital-based | Nigeria | None | Ekenze OS et al | Nigerian Journal of Medicine  NA | Not reported |
| 140 | The prevalence of neurological disorders in older people in Tanzania (2012). | Epidemiology | Cross-sectional  Population-based | Tanzania | UK | Dewhurst F et al | Acta Neurologica Scandinavica  3.2 | Research fellowship from the Dunhill Foundation and the Royal College of Physicians |
| 141 | Door-to-door survey of major neurological disorders (project) in Al Quseir City, Red Sea Governorate, Egypt (2013). | Epidemiology | Prospective Population-based | Egypt | None | El-Tallawy HN et al | Neuropsychiatric Disease and Treatment  2.9 | Not Reported |
| 142 | Prevalence of Parkinson’s disease and other types of Parkinsonism in Al Kharga district, Egypt (2013) | Epidemiology | Cross-sectional  Population-based | Egypt | None | El-Tallawy HN et al | Neuropsychiatric Disease and Treatment  2.9 | Not Reported |

Table 1(contd): Summary of included studies

| Study ID | Title and year | Theme | Design of study | African country(ies) defined by PD Cohort | Collaborating countries | First Author’s name | Journal and impact factor  2022 | Source of funding |
| --- | --- | --- | --- | --- | --- | --- | --- | --- |
| 143 | Prevalence of Parkinsonism and Parkinson’s disease in Qena governorate/Egypt: a cross-sectional community-based survey (2015). | Epidemiology | Cross-sectional  Population-based | Egypt | None | Khedr et al | Neurological Reearch  2.4 | Not reported |
| 144 | The pattern of neurological diseases in elderly people in outpatient consultations in Sub-Saharan Africa (2015) | Epidemiology | Retrospective Hospital-based | Cameroon | France | Callixte et al | BMC Research Notes  2.1 | Not reported |
| 145 | The Burden of Movement Disorders in Cameroon: A Rural and Urban-Based Inpatient/Outpatient Study (2017). | Epidemiology | Retrospective Hospital-based | Cameroon | Spain  USA | Cubo et al | Movement Disorders Clinical Practice  1.9 | World Federation of Neurology and the International Parkinson’s Disease and Movement Disorder Society |
| 146 | Burden of adult neurological diseases in Odeda Area, Southwest Nigeria (2020). | Epidemiology | Cross-sectional | Nigeria | None | Otubogun et al | BMJ Neurology  1.4 | None |

Table 1(contd): Summary of included studies

| Study ID | Title and year | Theme | Design of study | African country(ies) defined by PD Cohort | Collaborating countries | First Author’s name | Journal and impact factor  2022 | Source of funding |
| --- | --- | --- | --- | --- | --- | --- | --- | --- |
| 147 | Factors influencing the development of early- or late-onset Parkinson’s disease in a cohort of South African patients (2012). | Risk factors | Cross-sectional | South Africa | None | van der Merwe et al | South African Medical Journal  1.3 | Medical Research Council of SA, the Harry and Doris Crossley Foundation, and the University of Stellenbosch. |
| 148 | Neurologic Manifestation associated with an Outbreak of Typhoid fever, Malawi-Mozambique, 2009: An Epidemiologic Investigation (2012) | Risk factors | Retrospective  Cross-sectional | Malawi Mozambique | USA | Sejvar et al | PLoS One  3.2 | U.S. government funding, Centers for Disease Control and Prevention/Agency for Toxic Substances and Disease Registry. |
| 149 | Nutritional status and dietary habits in Parkinson’s disease patients in Ghana (2013). | Risk factors | Case- control study | Ghana | Italy | Barichella et al | Nutrition  3.1 | Fondazione Grigioni per il Parkinson |
| 150 | Trace Metals in Patients with Parkinson’s Disease: A Multi-Center Case-Control Study of Nigerian Patients (2013). | Risk factors | Cross sectional, Case-control | Nigeria | None | Ogunrin et al | Journal of Neurology and Epidemiology  NA | None |

Table 1(contd): Summary of included studies

| Study ID | Title and year | Theme | Design of study | African country(ies) defined by PD Cohort | Collaborating countries | First Author’s name | Journal and impact factor  2022 | Source of funding |
| --- | --- | --- | --- | --- | --- | --- | --- | --- |
| 151 | A Study of Extrapyramidal Manifestations Accompanying Decompensated Viral Hepatic Cirrhosis Patients (2017). | Risk factors | Cohort | Egypt | None | Ashour et al | Reviews on Recent Clinical Trials  1.4 | ASRT, Egypt |
| 152 | K-variant BCHE and pesticide exposure: Gene-environment interactions in a case–control study of Parkinson’s disease in Egypt (2018) | Risk factors | Retrospective  Cross-sectional | Egypt | Germany | Rösler et al | Scientific Reports  4.9 | German Academic Exchange Service (DAAD) within the Transformation Partnership Programme “Al Tawasul”. Günter Höglinger was funded by the Deutsche Forschungsgemeinschaf (DFG, HO2402/6-2 & Munich Cluster for Systems Neurology SyNergy), the German Federal Ministry of Education and Research (BMBF, 01KU1403A EpiPD), the Bavarian Ministry for Education, Culture, Science and Arts (Grant 8810001412 ForIPS), and the NOMIS foundation (FTLD project). |

Table 1(contd): Summary of included studies

| Study ID | Title and year | Theme | Design of study | African country(ies) defined by PD Cohort | Collaborating countries | First Author’s name | Journal and impact factor  2022 | Source of funding |
| --- | --- | --- | --- | --- | --- | --- | --- | --- |
| 153 | Manganese exposure, parkinsonian signs, and quality of life in South African mine workers (2019). | Risk factors | Cross-sectional | South Africa | UK  USA | Dlamini et al | American Journal of Industrial Medicine  2.2 | National Institute of Environmental Health Sciences (Grants: R01ES026891, K24ES017765, K01ES028295, and R21ES017504); American Parkinson Disease Association; and the Association of Commonwealth Universities. |
| 154 | GC-MS Based Metabolic Profiling of Parkinson’s Disease with Glutathione S-transferase M1 and T1 Polymorphism in Tunisian Patients (2020) | Risk factors | Cohort | Tunisia | Turkey | Rebai et al. | Combinatorial Chemistry & High Throughput Screening  1.3 | Not reported |
| 155 | Parkinsonism and chronic manganese exposure: Pilot study with clinical, environmental and experimental evidence (2020). | Risk factors | Prospective | Morocco | France | Kissani et al | Clinical Parkinsonism & Related Disorders  NA | None |

Table 1(contd): Summary of included studies

| Study ID | Title and year | Theme | Design of study | African country(ies) defined by PD Cohort | Collaborating countries | First Author’s name | Journal and impact factor  2022 | Source of funding |
| --- | --- | --- | --- | --- | --- | --- | --- | --- |
| 156 | Effects of glutathione S-transferase M1 and T1 deletions on Parkinson's disease risk among a North African population (2020). | Risk factors | Case-control | Tunisia | None | Rebai et al. | Revue Neurologique  2.6 | Not reported |
| 157 | Severity of parkinsonism associated with environmental manganese exposure (2021) | Risk factors | Retrospective  Cross-sectional | South Africa | UK  USA | Racette et al. | Environmental Health  7.1 | National Institute of Environmental Health Sciences, National Institute of Occupational Safety and Health (R01OH011661), Cure Alzheimer’s Fund, Dept of Defense (PD190057), Hope Center for Neurologic Disorders (Washington University). |
| 158 | Gut microbiota in Parkinson’s disease  patients: hospital‑based study (2021) | Risk factors | Cross-sectional  Case-control | Egypt | None | Khedr et al | The Egyptian Journal of Neurology,  Psychiatry and Neurosurgery  0.7 | Note reported |

Table 1(contd): Summary of included studies

| Study ID | Title and year | Theme | Design of study | African country(ies) defined by PD Cohort | Collaborating countries | First Author’s name | Journal and impact factor  2022 | Source of funding |
| --- | --- | --- | --- | --- | --- | --- | --- | --- |
| 159 | Relation of serum level of tumor necrosis  factor‑alpha to cognitive functions in patients  with Parkinson’s disease (2022). | Risk factors/  Biomarkers | Case-control | Egypt | None | El-Kattan et al. | The Egyptian Journal of Neurology,  Psychiatry and Neurosurgery  0.7 | None |
| 160 | Cytokines, miRNAs, and Antioxidants as Combined Non‑invasive  Biomarkers for Parkinson’s Disease (2022) | Risk factors/  Biomarkers | Cross-sectional  Case- control | Egypt | Italy | Ghit et al. | Journal of Molecular Neuroscience  3.4 | Not reported |
| 161 | Association of serum uric acid and non-motor symptoms in Parkinson's disease: A cross-sectional study from a movement disorders clinic in Lagos, Nigeria (2022) | Risk factors/  Biomarkers | Cross-sectional | Nigeria | None | Odeniyi et al | Journal of clinical sciences  6.1 | None |
| 162 | Risk Factors of Parkinson’s Disease: A Case-Control Study in Moroccan  Patients (2022) | Risk factors | Case-control | Morocco | None | Achbani et al | Arch Neuroscience  0.5 | None |

Table 1(contd): Summary of included studies

| Study ID | Title and year | Theme | Design of study | African country(ies) defined by PD Cohort | Collaborating countries | First Author’s name | Journal and impact factor  2022 | Source of funding |
| --- | --- | --- | --- | --- | --- | --- | --- | --- |
| 163 | Three years follow up for Levo-dopa plus Carbidopa treatment in a prevalent cohort of patients with Parkinson's disease in Hai, Tanzania (2011). | Management and Access to care | Prospective | Tanzania | None | Dotchin et al. | Journal of Neurology  4.8 | UK Parkinson's disease Society |
| 164 | The accessibility of Parkinson's disease medications in Kenya- Results of a national survey (2016). | Management and Access to care | Cross-sectional | Kenya | UK | Mokoya et al. | Movement disorder clinical practice  1.84 | African Task force of International Parkinson’s and Movement disorder society. |
| 165 | A nationwide survey of Parkinson's disease medicines availability and affordability in Nigeria (2019). | Management and Access to care | Survey | Nigeria | None | Okubadejo et al | Movement disorder clinical practice  1.84 | International Parkinson’s and Movement disorder society. |
| 166 | Availability of Therapies and Services for Parkinson's Disease in Africa: A Continent-Wide Survey (2021). | Management and Access to care | Survey | Egypt,Congo, Tunisia,Nigeria, Ethiopia,Cameroon, Morocco,Sudan, Algeria,Madagascar, Ivory Coast, Niger, Burkina Faso, Uganda, Somalia, Malawi, Burundi, Namibia, Libya, | Botswana, Chad, Djibouti, Zimbabwe, Kenya, Tanzania, Ghana, Zambia, South Africa | Hamid et al. | Movement Disorders  9.9 | Not reported |

Table 1(contd): Summary of included studies

| Study ID | Title and year | Theme | Design of study | African country(ies) defined by PD Cohort | Collaborating countries | First Author’s name | Journal and impact factor  2022 | Source of funding |
| --- | --- | --- | --- | --- | --- | --- | --- | --- |
| 167 | The effect of Origanum majorana tea on motor and non-motor symptoms in patients with idiopathic Parkinson's disease: a randomized controlled pilot study (2021). | Management and Access to care | A randomized double-blind placebo-controlled clinical trial | Tunisia | None | Chahra et al. | Parkinsonism  and related  disorders  4.8 | Tunisian Ministry of Higher Education and Scientific Research, and from the faculty  of Medicine Ibn el Jazzar, Sousse, Tunisia. |
| 168 | "We call the shaking illness': perception and experiences of PD in rural northern Tanzania (2011) | Patient engagement and QOL | Cross-sectional/  Survey | Tanzania | UK | Mshana G et al | BMC Public Health  3.9 | UK Parkinson’s disease society. |
| 169 | Profile of Generic and Disease-Specific Health-Related Quality (2014). | Patient engagement and QOL | Cross-sectional  Case-control | Nigeria | None | Okunoye et al | The Nigerian Health Journal  NA | None |
| 170 | Determinants of health-related quality of life among Nigerian-Africans with Parkinson’s disease (2014). | Patient engagement and QOL | Cross-sectional | Nigeria | None | Okunoye et al. | Port Harcourt Medical Journal  NA | None |

Table 1(contd): Summary of included studies

| Study ID | Title and year | Theme | Design of study | African country(ies) defined by PD Cohort | Collaborating countries | First Author’s name | Journal and impact factor  2022 | Source of funding |
| --- | --- | --- | --- | --- | --- | --- | --- | --- |
| 171 | Knowledge and Attitudes of Parkinson’s Disease in Rural and Urban Mukono District, Uganda: A Cross-Sectional, Community-Based Study (2015). | Patient engagement and QOL | Cross-sectional | Uganda | USA | Kaddumukasa et al | Parkinson's disease  5.5 | National Institute of Neurological Disorders and Stroke of the National Institute of Health under MEPI-Neurology Linked Award no. R25NS080968 |
| 172 | Beliefs, Knowledge and Attitudes Towards Parkinson's Disease Among a Xhosa Speaking Black Population in South Africa: A cross-sectional study | Patient engagement and QOL | Cross-sectional | South Africa | UK | Mokaya et al | Parkinsonism  and related  disorders  4.8 | Mandela Rhodes Foundation |
| 173 | Understanding the Experience and Perspectives of Parkinson's Disease Patients' Caregivers (2019). | Patient engagement and QOL | Cross-sectional | Ethiopia | None | Walga TK et al | Rehabilitation Research and Practice  0.5 | None |

Table 1(contd): Summary of included studies

| Study ID | Title and year | Theme | Design of study | African country(ies) defined by PD Cohort | Collaborating countries | First Author’s name | Journal and impact factor  2022 | Source of funding |
| --- | --- | --- | --- | --- | --- | --- | --- | --- |
| 174 | Efficacy of Cognitive Rehabilitation on Functional  Outcomes & Quality of Life in Parkinson’s Patients (2020). | Patient engagement and QOL | Cohort | Egypt | None | Elshamy et al | International journal of pharmaceutical research  0.03 | Not reported |
| 175 | Sexual dysfunction in male patients with Parkinson’s disease: related factors and impact on quality of life | Patient engagement and QOL | Cross-sectional | Egypt | None | Shalash et al | Neurological Sciences  3.8 | None |
| 176 | Mental Health, Physical Activity, and Quality of Life in Parkinson's Disease During COVID-19 Pandemic (2020) | Patient engagement and QOL | Case-control | Egypt | None | Shalash et al | Movement Disorders  9.9 | None |
| 177 | The role of support groups in the management of Parkinson’s disease in Kenya: Sociality, information, and legitimacy (2021). | Patient engagement and QOL | Interviews | Kenya | UK | Fothergill-Misbah et al. | Global Public Health  2.3 | This work was supported by Economic and Social Research Council: [Grant Number ES/J500082/1]. |

Table 1(contd): Summary of included studies

| Study ID | Title and year | Theme | Design of study | African country(ies) defined by PD Cohort | Collaborating countries | First Author’s name | Journal and impact factor  2022 | Source of funding |
| --- | --- | --- | --- | --- | --- | --- | --- | --- |
| 178 | Yield of training exchanges between Europe and Sub-Saharan Africa (2013). | Education and Training | Cohort | Cameroon | Belgium | Naeije G et al | Acta Neurol Belg  1.38 | None |
| 179 | A Parkinson's disease tele-education program for health care providers in Cameroon (2015). | Education and Training | quasi-experimental (comparison of 1 year baseline and post-education program intervention), hospital-based | Cameroon | Spain | Cubo E et al | Journal of the Neurological Sciences  3.1 | None |
| 180 | Telemedicine Enables Broader Access to Movement Disorders Curricula for Medical Students (2017) | Education and Training | quasi-experimental (comparison of 3months baseline and post-education program intervention), medical school based | Egypt | Spain  Argentina  Canada | Cubo et al | Tremor and Other Hyperkinetic Movements  1.6 | The Telemedicine Task Force of the International Parkinson’s disease and Movement Disorder Society |
