## Supplemental Materials for "The State of Play of Parkinson’s Disease in Africa: A Systematic Review and Point of View"

**Supplemental Table 2: Funding and Collaboration by African Regions and Countries**

| **Country** | **Collaboration** | **Funding** | |
| --- | --- | --- | --- |
| **North Africa** |  | **International** | **Local** |
| Tunisia | Turkey, France, Luxemburg, Germany, USA, Canada, Norway, UK, Taiwan, Poland, Switzerland, Japan | NIH grant, Europena Union Grant, Institut National de la Sante et de la Recherche Me´dicale GlaxoSmithKline, Mayo Foundation,  Michael J Fox Foundation for Parkinson’s Research, National Institute of Neurological Disorders and Stroke, Swiss National Science Foundation, Canada excellence research chairs program, the Cundill Foundation, province of British Columbia, Movement Disorder Society, German Research Foundation, the BMBF, the European Community | Institute of NeuologyTunis, Tunisian Ministry of Higher Education and Scientific Research, the Tunisian Ministry of Higher Education and Scientific Research, |
| Algeria | France | ANR and INSERM | Ministère de la Santé, de la Population et de la Réforme Hospitalière and the Ministère de l’Enseignement Supérieur et de la Recherche Scientifique |
| Egypt | Germany | The German Academic Exchange Service (DAAD) within the Transformation Partnership Programme the Deutsche Forschungsgemeinschaf (DFG, HO2402/6-2 & Munich Cluster for Systems Neurology SyNergy), the German Federal Ministry of Education and Research, the Bavarian Ministry for Education, Culture, Science and Arts and the NOMIS foundation (FTLD project). | ASRT, Egypt |
| Libya | India |  |  |
| Morocco | France, | Novartis Pharma, | Centre National de Recherche Scientifique et Technique” (CNRST), Minist`ere de l’Enseignement Sup´erieur, de la Recherche Scientifique et de la Formation des Cadres” (MESRSFC), Mohammed V University in Rabat (UM5R), Laboratory of Cell Biology and Molecular Genetics, Faculty of Sciences of Agadir |
| **East Africa** |  |  |  |
| Sudan | France, Saudi Arabia, | NIH grant |  |
| Tanzania | UK, Netherlands | UK Parkinson’s Disease Society and Northumbria Healthcare NHS Trust, British Academy UK-Africa Academic Partnerships, Dunhill Foundation, the Royal College of Physicians, the Peel Medical Research Trust, a British Geriatric Society SpR start-up grant, an Academy of Medical Sciences (UK) Clinical Lecturer start-up grant and Northumbria Healthcare NHS Foundation Trust. |  |
| Kenya | UK | The African TaskForce of International Parkinon's and Movement disorder society, Economic and Social Research Council, |  |
| Uganda | USA | National Institute of Neurological Disorders and Stroke of the National Institute of Health under MEPI-Neurology Linked Award |  |
| Ethiopia | Sweden, USA | Swedish Agency for Research Cooperation with Developing Countries | Addis Ababa University |
| Madagascar |  |  |  |
| **West Africa** |  |  |  |
| Nigeria | UK, USA, Portugal | NIH, NIA grant, American Parkinson Disease Association (APDA) Mayo Clinic Information and Referral Center/ Polish National Agency for Academic Exchange Iwanowska's Fellowship/ National Institutes of Health (NIH)/National Institute of Neurological Disorders and Stroke (NINDS), the University College London Global Challenge Research Grant, International Parkinon's and Movement disorder society, | University of Lagos Central Research Committee Grant |
| Ghana | UK, USA | Fondazione Grigioni per il Morbo di Parkinson and Regione Lombardia |  |
| Senegal | France |  |  |
| **South Africa** |  |  |  |
| South Africa | UK, USA, Netherlands, Canada, France, | Agence Nationale de la Recherche, Michael J Fox Foundation, mayo clinic, NIH, the American Parkinson Disease Association (APDA), the National Institutes of Health–National Institute of Environmental Health Sciences, American Parkinson Disease Association; and the Association of Commonwealth Universities. | South African Medical Research Council, the Harry and Doris Crossley Foundation and the University of Stellenbosch, NRF-DST Centre of Excellence for Biomedical Tuberculosis Research, the National Research Foundation of South Africa, The Nelson Mandela Metropolitan University (NMMU) and the National Research Foundation (NRF), University of the Witwatersrand School of Public Health, Mandela Rhodes Foundation, |
| Zambia | Netherlands | International Parkinson fonds, the Netherlands and Netherlands Organisation for Scientific Research |  |
| Malawi/Mozambique | USA | U.S. government funding, Centers for Disease Control and Prevention/Agency for Toxic Substances and Disease  Registry. |  |
| **Central Africa** |  |  |  |
| Cameroon | Spain, Argentina, Canada, Belgium, USA, France, Australia, | Research Intramural Program of the Carlos III Institute of Health, World Federation of Neurology and the International Parkinson’s Disease and Movement Disorder Society, telemedicine Task Force of the International Parkinson’s Disease and Movement Disorder Society |  |
| **Inter African collaboration** |  |  |  |
| Algeria, Morocco & Tunisia | France, USA, JAPAN, UK, Portugal |  |  |
| Algeria, Morocco, Tunisia & Libya | Turkey, Japan, Algeria, South Africa, France | Agence Nationale de la Recherche |  |
| South Africa & Nigeria | USA | NINDS, NIH Fogarty | South African National Research Foundation, South African Medical Research Council. Faculty of Medicine and Health Sciences, Stellenbosch University, South Africa. South African Tuberculosis Bioinformatics Initiative (SATBBI), |
| Nigeria, Cameroon & Egypt | USA, Spain |  |  |
| Ghana & Zambia | Italy | ‘Fondazione Grigioni per ilMorbo di Parkinson’, Milan, Italy |  |
| Egypt, Congo, Tunisia, Nigeria, Ethiopia, Cameroon, Morocco, Sudan, Algeria, Madagascar, Ivory Coast, Niger, Burkina Faso, Uganda, Somalia, Malawi, Burundi, Namibia, Libya, Botswana, Chad, Djibouti, Zimbabwe, Kenya, Tanzania, Ghana, Zambia, South Africa | NA | NA |  |

**Supplemental material 1**

**Search terms**

The word “Parkinson” OR “Parkinsonian disorder” (Medical Subject Heading (MESH

or text word) were combined with “Africa” and each “African country:

(“Algeria” OR “Angola”) OR (“Botswana” OR “Burundi” OR “Burkina Faso” OR “Benin”) OR (“Comoros” OR “Cabo Verde” OR “Congo” OR “Central African Republic” OR “Chad” OR “Cameroon” OR “Cote d'Ivoire" OR "Ivory Coast") OR (“Djibouti” OR “Democratic Republic of Congo” OR “DR Congo”) OR (“Eswatini” OR “Eritrea” OR “Equatorial Guinea” OR “Ethiopia” OR “Egypt”) OR (“Gambia” OR “Gabon” OR “Guinea-Bissau” OR “Ghana” OR “Guinea”) OR (“Kenya”) OR (“Libya” OR “Liberia” OR “Lesotho”) OR (“Mauritania” OR “Mauritius” OR “Mali” OR “Malawi” OR “Madagascar” OR "Morocco” OR “Mozambique”) OR (“Namibia” OR ““Niger” OR “Nigeria”) OR (“Rwanda”) OR (“Sao Tome & Principe” OR “Seychelles” OR “South Sudan” OR “Sierra Leone” OR “Senegal” OR “Somalia” OR “Sudan” OR “South Africa”) OR (“Tunisia” OR “Togo” OR Tanzania”) OR (“Zambia” OR “Zimbabwe”) OR (“Uganda”) OR (“sub-Saharan Africa” OR “Africa South of the Sahara”).

**Inclusion and Exclusion criteria**

Original studies with any design were considered potentially eligible if the participants included were African, had PD, and if the studies provided information on any of the focus areas including epidemiology, genetics, risk factors, clinical characteristics, patient-reported outcomes (experience and quality of life), disease management and outcomes, access to care, patient support, and healthcare workforce training. There were no restrictions with regard to PD diagnostic criteria, language, or year of publication. Expert opinions, some letters to the editor, case reports, editorials, and reviews were excluded.

**Study Selection**

Two of the investigators searched independently (OO and AC) while five of the investigators (OO, AC, YZ, BA, SD) did the first study selection of original articles independently by going through the search results according to the title or quick review of abstracts. The selected full articles were independently read and screened for eligibility by two reviewers (JA, MA). To identify other relevant studies, two teams members (YZ, OO) conducted forwards and backward citation tracking of key articles. Disagreements were resolved through discussion with the whole team.

**Data extraction**

A standardized data extraction form was used to record data of interest such as the socio-demographic characteristics of the study, the record of information on all authors, cohort/study name, year of publication, country, study design, study population, the proportion of males and females, mean age of subjects/controls, number of people with PD, type of statistical analysis, information on epidemiology, information on clinical features, data on risk factors for PD including genetics, environmental risk factors such as diet, viruses, infections, access to care, information on available medications, data on quality of life and patients experiences including stigma and cultural beliefs, data on patients support groups and organizations, training and educational resources/active organizations, funding and research output and data on recommendations for future studies (Appendix 2).

**Methodological quality**

The Newcastle-Ottawa scale was used to conduct the quality appraisal of included studies.^24^ This tool examines whether studies demonstrated in clear terms the method of selection of the population of interest, quantitative methodology, accuracy of the recruitment process, method of data collection and evaluation of outcome. Studies are likely to have overall scores ranging from 0 to 9, with scores ranging from 0 to 3 defining low-quality studies, 4 to 6 defining medium-quality studies and 7 to 9 defining high-quality studies. The Joanna Briggs Institute checklist (JBI) was used to score three articles where the Newcastle-Ottawa scale was not suitable.

**Appendix 1: Medline Search**

MEDLINE Search

| 1. exp Africa/ |
| --- |
| 2. africa, eastern/ or burundi/ or djibouti/ or comoros/ or eritrea/ or ethiopia/ or kenya/ or rwanda/ or somalia/ or south sudan/ or sudan/ or tanzania/ or uganda/ |
| 3. africa, northern/ or algeria/ or egypt/ or libya/ or morocco/ or tunisia/ |
| 4. "africa south of the sahara"/ or africa, central/ or africa, eastern/ or africa, southern/ or africa, western/ |
| 5. africa, western/ or benin/ or burkina faso/ or cabo verde/ or cote d'ivoire/ or ivory coast/ or gambia/ or ghana/ or guinea/ or guinea-bissau/ or liberia/ or mali/ or mauritania/ or mauritius/ or niger/ or nigeria/ or senegal/ or sierra leone/ or togo/ |
| 6. africa, central/ or cameroon/ or central african republic/ or chad/ or congo/ or "democratic republic of the congo"/ or equatorial guinea/ or gabon/ or "sao tome and principe"/ or seychelles.mp. [mp=title, abstract, original title, name of substance word, subject heading word, floating sub-heading word, keyword heading word, organism supplementary concept word, protocol supplementary concept word, rare disease supplementary concept word, unique identifier, synonyms] |
| 7. africa, southern/ or angola/ or botswana/ or eswatini/ or lesotho/ or malawi/ or mozambique/ or namibia/ or south africa/ or zambia/ or zimbabwe/ or sub-Saharan Africa.mp. [mp=title, abstract, original title, name of substance word, subject heading word, floating sub-heading word, keyword heading word, organism supplementary concept word, protocol supplementary concept word, rare disease supplementary concept word, unique identifier, synonyms] |
| 8. africa*.tw. |
| 9. 1 or 2 or 3 or 4 or 5 or 6 or 7 or 8 |
| 10. exp Parkinsonian Disorders/ |
| 11. parkinson*.tw. |
| 12. 10 or 11 |
| 13. 9 and 12 |

**Appendix 2: Data Collection Form**

**Date:**

**Version No:**

**Title of review: The State of Play of Parkinson’s Disease (PD) in Africa: A systematic review and a Point of View**

**Sub-title:**

**The number of articles with data on sub-title:**

A. **Identification**

Review Author ID/Name:

Study ID (Name and year):

Report ID:

Citation and contact details:

B. **Eligibility**

**Criteria for Eligibility	Yes	No**

Original article of African PD cases

Reports any information on PD in Africa

**Reasons for exclusion	Yes	No**

Data not about PD cases in Africa

Failed quality assessment

C. **Methods	Yes	No**

**Study Design**

Prospective cohort study

Retrospective cohort study

Case-control study

Cross-sectional study

Case report/case series

**Study duration (years)**

<1

1-5

>5

**Subjects**

Total number of PD patients

Total number of controls (if available)

Mean Age

Sex

Country

Population: Hospital-based

Population-based

D. **Results**
E. **Miscellaneous**

Key conclusion of the study

References to other relevant studies

Correspondence required
